# Transcriptomic Response of Postprandial Blood, Subcutaneous Adipose Tissue and Muscle to a combined lifestyle intervention in older adults

**DOI:** 10.1101/2023.08.04.23293606

**Authors:** F.A. Bogaards, T. Gehrmann, M. Beekman, N. Lakenberg, E. Suchiman, C.P.G.M. de Groot, M.J.T. Reinders, P.E. Slagboom

**Affiliations:** Molecular Epidemiology, Leiden University Medical Center, Leiden, The Netherlands; Leiden Computational Biology Center, Leiden, The Netherlands; Human Nutrition and Health, Wageningen University & Research, Division of Human Nutrition, Wageningen, The Netherlands; Delft Bioinformatics Lab, Delft University of Technology, Delft, The Netherlands; Max Planck Institute for Biology of Aging, Cologne, Germany

## Abstract

Molecular effects of lifestyle interventions are typically studied in a single tissue. Here, we investigated the sex-specific effects of the Growing Old TOgether (GOTO) study, a moderate 13-week combined lifestyle intervention on the transcriptomes of postprandial blood, subcutaneous adipose tissue (SAT) and muscle tissue in healthy older adults, the overlap in effect between tissues and their relation to whole-body parameters of metabolic health. The GOTO intervention had virtually no effect on the postprandial blood transcriptome, while the SAT and muscle transcriptomes responded significantly. In SAT, pathways involved in HDL remodeling, O_2_/CO_2_ exchange and signaling were overrepresented, while in muscle, collagen and extracellular matrix pathways were significantly overexpressed. Additionally, we found that the effects of the SAT transcriptome closest associated with gains in metabolic health. Lastly, in males, we identified a shared variation between the transcriptomes of the three tissues. We conclude that the GOTO intervention had a significant effect on metabolic and muscle fibre pathways in the SAT and muscle transcriptome, respectively. Aligning the response in the three tissues revealed a blood transcriptome component which may act as an integrated health marker for metabolic intervention effects across tissues.

## Main

Worldwide, the proportion of older people in our society is increasing, and with it the burden of age-related diseases on the quality of life and health economics ^1,2^. Improving metabolic health is considered key to improving vitality at older ages ^3,4^. Dietary, activity-based, or combined lifestyle interventions are known to improve culprit factors in human ageing such as metabolic health and immunity. Although effects are usually heterogeneous, the metabolic health can significantly be improved, even in middle and older age ^5–10^.

The effect of lifestyle interventions can be studied by traditional blood-based markers of metabolic health (including: BMI, fasting insulin, systolic blood pressure (SBP), HDL cholesterol, diastolic blood pressure (DBP), dual-energy X-ray absorptiometry (DEXA)-based whole-body fat%, trunk fat%) ^6,11^ and a range of omics based assays such as the metabolome^6,12–14^, proteome ^15,16^, epigenome ^8^ and the transcriptome ^17–27^, of blood ^6,12–17,20,28,29^, gut ^15^, adipose tissue ^18,21,26,30,31^, and muscle tissue ^19,22,23,27^. Human intervention studies are usually small (n = 120 – 250) ^6,29,32–34^ and focus on one tissue only, mainly blood. Mostly, a strong effect is observed on the fasting blood proteome ^16^ and metabolome ^6^, but typically not on the fasting transcriptome, where the effects are milder and more variable ^17,24,35^, except for rigorous lifestyle interventions ^29^. The transcriptomes of adipose tissue and muscle do react respectively on separate dietary ^25^ and physical activity ^19,22,23,27^ lifestyle interventions. In mice, combined lifestyle interventions were shown to affect the transcriptome of brown adipose tissue and skeletal muscle ^36^.

In humans, however, the simultaneous effect of combined lifestyle interventions on the transcriptome of blood, subcutaneous adipose tissue (SAT) and muscle was not investigated as yet. To understand how differences in omics profiles relate to changes over the years in health parameters one often relies on population-based omics studies. However, such studies mainly rely on blood samples ^37,38^. We speculate that some of the transcriptomic changes in different tissues in response to interventions align due to the systemic nature of lifestyle interventions and that a part of changes in the blood transcriptome reflect responses in SAT and muscle and related health gains. Hereto, an integrative analysis of the SAT, muscle and blood transcriptomic response to such intervention is necessary.

Integrative data analysis methods pool data from different datasets to jointly analyze them ^39^. Joint and Independent Variation Explained (JIVE) ^40^ is one such integrative data analysis method. JIVE decomposes the variation across the different datasets in three components: 1) the joint variation between the datasets, 2) dataset-specific variations, and 3) the residual noise for each dataset separately. The blood transcriptome response in mild interventions is usually minimal, JIVE however may reveal a part that represents the joint response variation across tissues. We are interested whether the joint response captures the intervention effects on metabolic health stronger than the blood transcriptome alone.

We applied the tissue-specific as well as the integrative data analysis approach on a 13-week combined physical activity and dietary lifestyle intervention study in older adults; the Growing Old TOgether (GOTO) study. Within this study the transcriptome of postprandial blood, SAT and muscle tissue before and after intervention has been measured. We report on genes whose expressions within each of the different tissues associated with whole body health improvements across the intervention, and report on the observed joint transcriptional effect. We conclude that especially the SAT and muscle transcriptome respond to the intervention by HDL remodeling, cell signaling and CO_2_/O_2_ exchange for SAT, and collagen/extracellular matrix pathways for muscle. Especially the SAT transcriptome in both sexes associates to beneficial changes in whole-body composition. The blood transcriptome does not generally respond to intervention or reflect health gain. Our integrative analysis of blood, however, revealed that the joint effect of the three tissues associated with the parameters of whole-body composition parameters. This blood transcriptome component may act as an integrated health marker for metabolic intervention effects across tissues.

## Results

### Health markers improve but improvement varies among subsets

Participants compliant to the intervention overall improved their metabolic health (Table 1) with for some parameters slight sex-differences in magnitude of effect. We have transcriptome data available in subsets of the complete GOTO study group: 1) Overlapping Blood, SAT and Muscle 57, which consists of 33 males and 24 females, 2) Blood Postprandial Samples 88, which contain 45 males and 43 females, 3) SAT Postprandial samples 78, which contains 38 males and 40 females, and 4) Muscle Postprandial samples 82, which contains, 48 males and 34 females. The direction of effect on the health parameters in the subsets of participants for which data in different tissues is available was similar (Table 1), though there were some differences in significance level. For example, in males, the intervention only had a significant effect on HDL cholesterol in the group with muscle transcriptome measurements. While in females, HDL cholesterol size was significantly different in only the participants with SAT transcriptome measurements, and the FRS was significantly different in the complete set of measurements. These differences in effect could be partially explained by slight baseline classical metabolic health marker level differences between the groups (Supplementary Table 1). To circumvent the slight differences in intervention effect on the classical metabolic health markers between the different groups, the associations between the transcriptome and the classical metabolic health markers will be only be calculated on the overlapping postprandial blood, SAT and muscle samples.

**Table 1:** Effect of the 13-weeks lifestyle intervention on metabolic health marker measurements differs per subset selection.

| Health Marker, mean (SD) | Sex | Complete GOTO dataset<br>153<br>(75 Male, 78 Female) |  | Overlapping Blood, SAT and<br>Muscle<br>Samples 57<br>(33 Male, 24 Female) |  | Blood Postprandial Samples<br>88 (45 Male, 43 Female) |  | SAT Postprandial Samples<br>78 (38 Male, 40 Female) |  | Muscle Postprandial<br>Samples<br>82 (48 Male, 34 Female) |  |
| --- | --- | --- | --- | --- | --- | --- | --- | --- | --- | --- | --- |
|  |  | Delta (sd) | P-adjust | Delta (sd) | P-adjust | Delta (sd) | P-adjust | Delta (sd) | P-adjust | Delta (sd) | P-adjust |
| BMI, kg/m<sup>2</sup> | Male | -1.09 (0.78) | 2.4e-18* | -1.02 (0.78) | 1.8e-07* | -1.15 (0.79) | 1.4e-11* | -1.10 (0.77) | 1.3e-09* | -1.18 (0.78) | 6.9e-13* |
| Waist Circumference | Male | -4.49 (5.26) | 1.8e-09* | -3.82 (4.97) | 1.1e-03* | -4.47 (5.29) | 1.1e-05* | -4.42 (5.10) | 4.8e-05* | -4.94 (5.41) | 8.7e-07* |
| Systolic Blood Pressure, mm Hg | Male | -3.25 (12.54) | 2.8e-01 | -3.80 (14.36) | 1.0e+00 | -2.52 (13.42) | 1.0e+00 | -3.91 (14.14) | 9.7e-01 | -3.05 (13.53) | 1.0e+00 |
| Diastolic Blood Pressure, mm Hg | Male | -0.75 (6.98) | 1.0e+00 | -3.04 (7.09) | 1.9e-01 | -2.27 (6.67) | 2.7e-01 | -3.09 (7.10) | 1.1e-01 | -2.32 (6.86) | 2.3e-01 |
| Fasting Insulin, mU/L | Male | -0.31 (3.30) | 1.0e+00 | 0.14 (3.63) | 1.0e+00 | 0.04 (3.62) | 1.0e+00 | -0.11 (3.71) | 1.0e+00 | -0.23 (3.72) | 1.0e+00 |
| Whole-Body Fat, % | Male | -1.60 (1.65) | 1.2e-09* | -1.17 (1.55) | 1.5e-03* | -1.43 (1.71) | 8.2e-06* | -0.92 (1.52) | 9.3e-05* | -1.77 (1.66) | 4.8e-07* |
| Trunk Fat, % | Male | -2.47 (2.13) | 3.2e-12* | -1.96 (1.81) | 3.3e-05* | -2.26 (2.13) | 2.3e-07* | -1.48 (1.74) | 1.1e-06* | -2.59 (2.04) | 8.7e-09* |
| Fasting HDL Cholesterol, mmol/L | Male | 0.04 (0.14) | 8.2e-02 | 0.04 (0.13) | 1.0e+00 | 0.06 (0.14) | 7.7e-02 | 0.05 (0.13) | 3.4e-01 | 0.07 (0.14) | 1.7e-02* |
| Fasting HDL Cholesterol size, nm | Male | 0.06 (0.09) | 2.0e-06* | 0.06 (0.08) | 9.9e-04* | 0.08 (0.08) | 2.0e-06* | 0.06 (0.09) | 4.6e-04* | 0.08 (0.09) | 1.3e-06* |
| FRS | Male | -0.40 (1.22) | 5.8e-02 | -0.61 (1.37) | 1.6e-01 | -0.53 (1.25) | 6.6e-02 | -0.58 (1.31) | 9.7e-02 | -0.54 (1.24) | 3.9e-02* |
| BMI, kg/m<sup>2</sup> | Female | -1.23 (0.82) | 1.4e-20* | -1.26 (0.72) | 1.4e-07* | -1.27 (0.64) | 2.7e-15* | -1.35 (0.73) | 3.0e-13* | -1.36 (0.74) | 2.5e-11* |
| Waist Circumference | Female | -4.33 (5.57) | 1.4e-08* | -4.71 (5.74) | 5.4e-03* | -4.70 (5.30) | 7.5e-06* | -4.58 (5.31) | 3.0e-05* | -5.21 (5.32) | 2.3e-05* |
| Systolic Blood Pressure, mm Hg | Female | -3.01 (11.69) | 2.6e-01 | -1.67 (11.69) | 1.0e+00 | -2.85 (11.30) | 1.0e+00 | -2.46 (11.73) | 1.0e+00 | -1.35 (12.07) | 1.0e+00 |
| Diastolic Blood Pressure, mm Hg | Female | -1.25 (6.52) | 9.5e-01 | -1.19 (5.75) | 1.0e+00 | -1.66 (5.64) | 6.1e-01 | -2.29 (6.27) | 2.6e-01 | -1.43 (6.54) | 1.0e+00 |
| Fasting Insulin, mU/L | Female | -0.36 (3.25) | 1.0e+00 | -0.30 (2.35) | 1.0e+00 | -0.02 (3.01) | 1.0e+00 | -0.33 (3.63) | 1.0e+00 | -0.90 (2.84) | 7.3e-01 |
| Whole-Body Fat, % | Female | -1.57 (1.70) | 9.8e-09* | -1.33 (2.22) | 1.1e-02* | -1.64 (1.85) | 5.2e-05* | -1.58 (1.98) | 7.2e-05* | -1.98 (2.07) | 2.5e-04* |
| Trunk Fat, % | Female | -2.22 (2.15) | 3.1e-10* | -1.76 (2.78) | 7.1e-03* | -2.32 (2.36) | 1.2e-05* | -2.31 (2.48) | 2.6e-05* | -2.60 (2.57) | 1.1e-04* |
| Fasting HDL Cholesterol, mmol/L | Female | -0.04 (0.15) | 2.7e-01 | -0.03 (0.14) | 1.0e+00 | -0.05 (0.16) | 4.7e-01 | -0.05 (0.15) | 5.9e-01 | 0.00 (0.14) | 1.0e+00 |
| Fasting HDL Cholesterol size, nm | Female | 0.03 (0.08) | 1.7e-02* | 0.05 (0.09) | 1.7e-01 | 0.02 (0.08) | 6.0e-01 | 0.04 (0.08) | 2.6e-02* | 0.04 (0.09) | 9.1e-02 |
| FRS | Female | -0.65 (1.60) | 5.5e-03* | -0.46 (1.59) | 1.0e+00 | -0.42 (1.47) | 6.8e-01 | -0.62 (1.76) | 3.1e-01 | -0.68 (1.79) | 3.4e-01 |

Abbreviations: SD; standard deviation, SAT; subcutaneous adipose tissue, BMI; body mass index, FRS; Framingham Risk Score. Associations were calculated using a liner mixed model, adjusting for age at baseline and person ID (as a random effect). P-values were adjusted using the Bonferroni correction method, an adjusted p-value < 0.05 was indicated with an \*.

### Strong sex-dependent transcriptional response in subcutaneous adipose tissue and muscle

We investigated the effect of the GOTO intervention by measuring transcriptional differences in blood, SAT and muscle (differential gene expression analysis, see Materials and Methods). In males, the number of significantly differentially expressed genes (DEGs) across the intervention varied considerably between blood (one), SAT (89), and muscle (251) (Figure 1, Table 2, Supplementary Table 2, 3 & 4). The ratio between up and down regulated DEGs also differed between the affected tissues: a slightly larger number of downregulated genes in SAT whereas over 80% of the DEGs were upregulated in muscle. Noteworthy, the muscle DEGs contained 14 collagen genes, all of which were upregulated (Supplementary Table 4).

**Figure 1:**
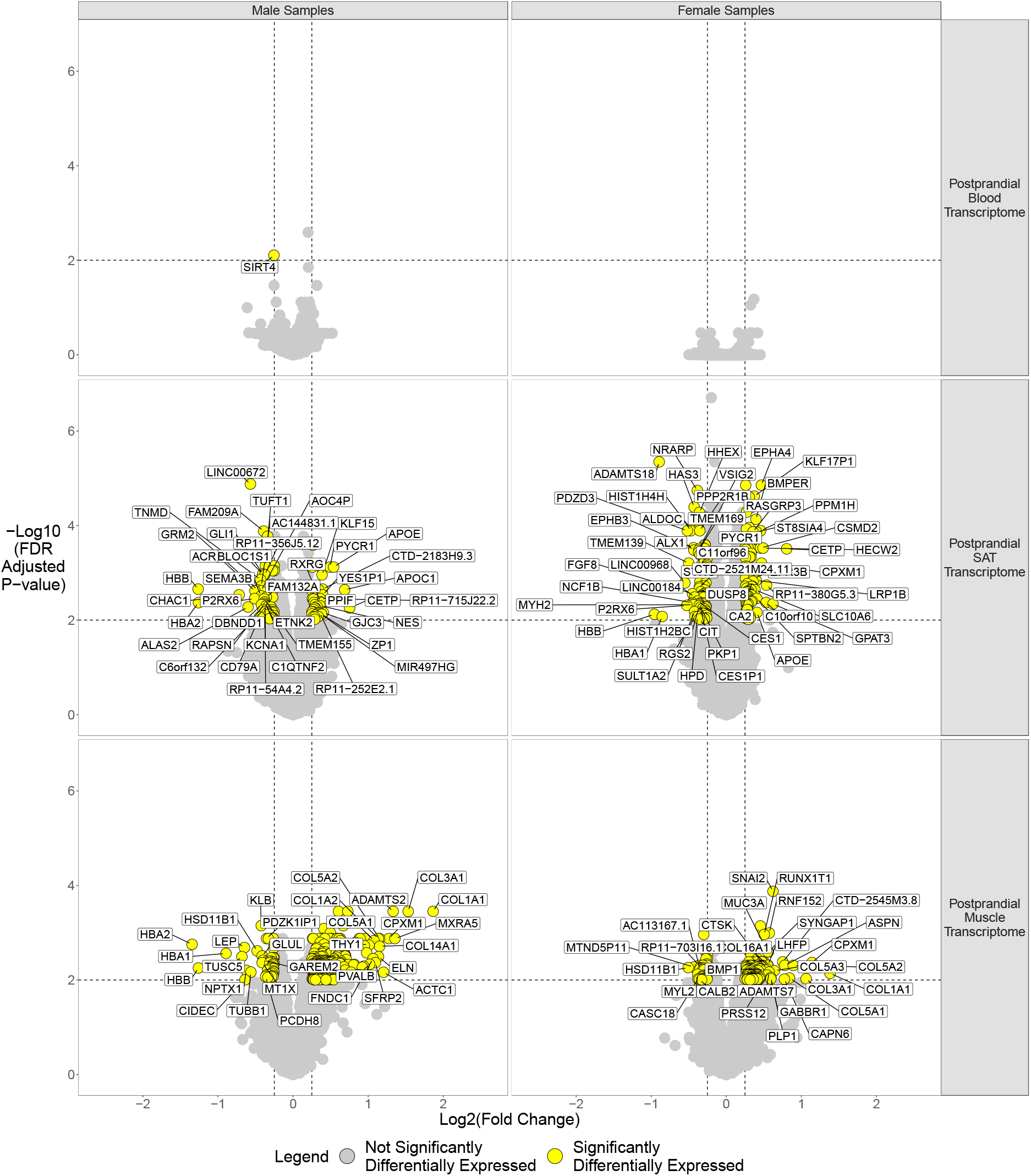
Intervention effect on postprandial blood, SAT and muscle gene expression levels in men and women. The log2 fold change is plotted on the x-axis, the -log10 FDR adjusted p-value is plotted on the y-axis. The dashed horizontal line represents the significance threshold of FDR 0.01, the two vertical lines represent the log2 fold change thresholds of -0.25 and 0.25. Genes with an absolute log2 fold change ≥ 0.25 and an FDR adjusted p-value < 0.01 were considered significantly differentially expressed. Each dot represents a gene. Yellow dots represent the significantly differentially expressed genes (DEGs). Grey dots represent genes with an absolute log2 fold change < 0.25 and/or FDR ≥ 0.01. Significantly differentially expressed genes with the strongest log2 fold changes were labeled with their gene symbol. Abbreviations: SAT; subcutaneous adipose tissue.

**Table 2:** The 13-weeks lifestyle intervention had the strongest effect on postprandial SAT and Muscle.

| Sex | Tissue | State | Direction of Effect | Expressed Genes | Genes with Log2FC ≥ 0.25 | Genes with FDR < 0.01 | Genes with Log2FC ≥ 0.25 & FDR < 0.01 |
| --- | --- | --- | --- | --- | --- | --- | --- |
| Male | Blood | Postprandial | Downregulated | 8602 | 52 | 1 | 1 |
| Male | Blood | Postprandial | Upregulated | 10093 | 45 | 1 | 0 |
| Male | SAT | Postprandial | Downregulated | 10099 | 402 | 123 | 48 |
| Male | SAT | Postprandial | Upregulated | 9614 | 173 | 180 | 41 |
| Male | Muscle | Postprandial | Downregulated | 9413 | 228 | 139 | 46 |
| Male | Muscle | Postprandial | Upregulated | 7166 | 674 | 239 | 205 |
| Female | Blood | Postprandial | Downregulated | 9619 | 63 | 0 | 0 |
| Female | Blood | Postprandial | Upregulated | 9143 | 32 | 0 | 0 |
| Female | SAT | Postprandial | Downregulated | 9860 | 254 | 331 | 92 |
| Female | SAT | Postprandial | Upregulated | 9599 | 143 | 267 | 76 |
| Female | Muscle | Postprandial | Downregulated | 9464 | 292 | 127 | 46 |
| Female | Muscle | Postprandial | Upregulated | 7570 | 827 | 132 | 122 |

*Abbreviations: SAT; subcutaneous adipose tissue*

For females, the number of DEGs across the different tissues were somewhat similar as in males, but the effect sizes were smaller (Figure 1, Table 2): zero DEGs in blood, 168 in SAT, and 168 in muscle (Supplementary Table 5 & 6). Like the male samples, more than half of the DEGs in SAT were downregulated, while in muscle over two-thirds of significant DEGs were upregulated. The female muscle DEGs contained 7 collagen genes, all of them upregulated (Supplementary Table 6).

Overall, in both males and females, the intervention had hardly any effect on the blood transcriptome while in SAT both up- and downregulated genes were observed, whereas in muscle the majority of the DEGs were upregulated.

### Lipid and collagen related pathways enriched in SAT and muscle response, respectively

We then investigated which pathways were associated with the found DEGs (pathway overrepresentation analysis, see Materials and Methods). In male SAT, five pathways were significantly overrepresented (Figure 2, Supplementary Table 7). The three upregulated pathways were involved with HDL remodeling, nuclear receptor signaling and cholesterol transport, while the downregulated pathway was involved in fertilization, the remaining pathway (equally down -and upregulated) was involved in ligand clearance. In male muscle, all 18 enriched pathways were upregulated. The majority of these were involved in collagen formation or extracellular matrix formation/degradation (Figure 2, Supplementary Table 7). The remaining pathways were involved in myogenesis (NCAM1 pathways), mesenchymal stem cell differentiation (PDGF pathway), scavenger receptors (binding and uptake, class A receptors) and protein O-glycosylation.

**Figure 2:**
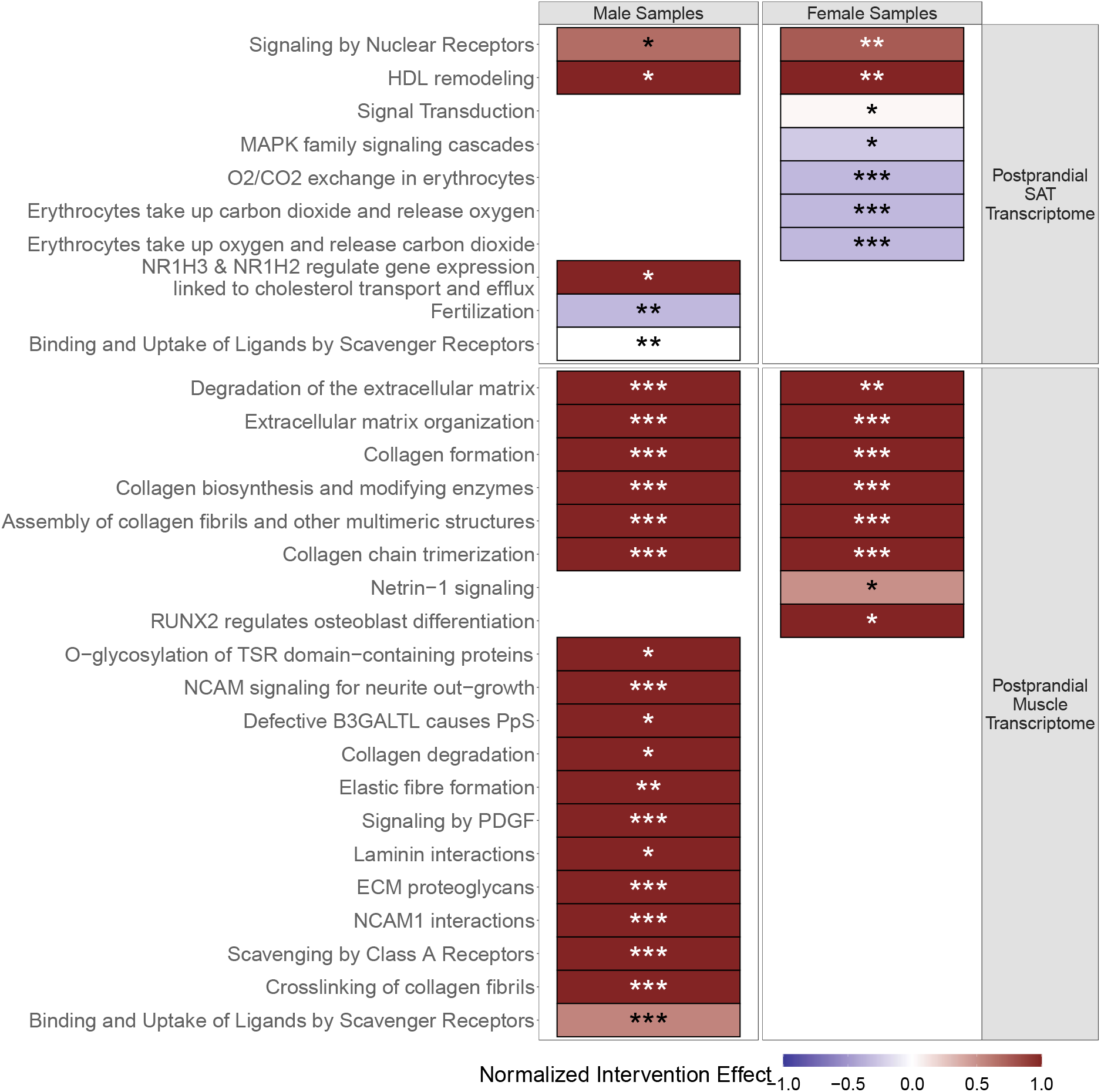
Significantly Overrepresented pathways in Postprandial SAT and Muscle Transcriptome. The overrepresented pathways are plotted on the y-axis. Overrepresented pathways in the male samples are plotted on the left, overrepresented samples in female samples, on the right. The fill color represents the normalized direction of effect: dark blue (-1) all significantly differentially expressed genes in the pathway were underexpressed, dark red (1) all significantly differentially expressed genes in the pathway were overexpressed. P- values were adjusted for multiple testing using the Bonferroni correction method. Significance is indicated by the asterisks (* = p- adjust <0.05, ** = p- adjust <0.01, *** = p- adjust <0.001). Abbreviations: SAT; subcutaneous adipose tissue.

In female SAT, seven pathways were significantly overrepresented (Figure 2, Supplementary Table 7). The three upregulated pathways were involved in signaling pathways and HDL remodeling. The four downregulated pathways were related to MAPK signaling and CO_2_/O_2_ exchange. However, the three CO_2_/O_2_ pathways were only overrepresented due to the same three genes (Supplementary Table 7). In female muscle, all eight overrepresented pathways were upregulated (Figure 2, Supplementary Table 7), six of which were involved in collagen formation and extracellular matrix organization/degradation. The remaining pathways were involved in in netrin-1 signaling and osteoblast differentiation.

Taken together, both in males and females, the overrepresented pathways in muscle were largely involved in collagen and extracellular matrix remodeling. In SAT, there were fewer enriched pathways which were less consistent in terms of direction (both -up and downregulated). The overrepresented pathways in SAT were involved in signaling, O_2_/CO_2_ exchange, HDL remodeling and cellular signaling.

### Response of tissue-specific DEGs reproduced across SAT and muscle, not in blood

Next, we questioned whether there was a transcription response common to the three tissues. Hereto, we examined the response of the eigengene of a set of tissue-specific DEGs in the other tissues (see Materials and Methods). For example, the eigengene of the male-upregulated DEGs in SAT tissue was also positively upregulated in muscle, whereas it was non-significantly downregulated in blood (second row in Figure 3). Similarly, in females, we noted correspondence between SAT and muscle, although the eigengene of the upregulated DEGs in SAT was stronger than in males, in blood the effect was less significant (sixth row, Figure 3). Since there was only one significant DEG in postprandial blood (Figure 1, Table 2), no blood down- or upregulated DEGs eigengenes were calculated.

**Figure 3:**
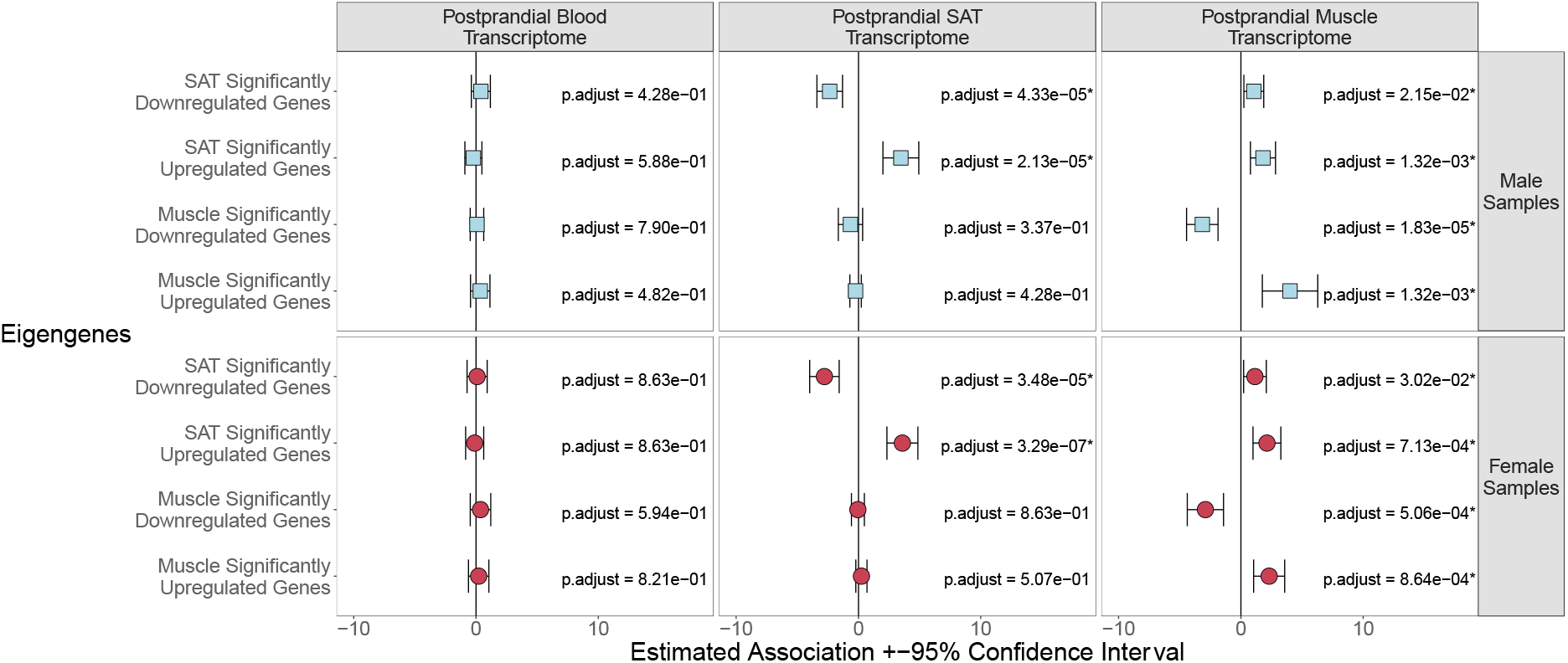
Effect of the 13-weeks lifestyle intervention on Eigengenes calculated using up- or downregulated significantly differentially expressed genes. Genes used for input of the eigengenes were plotted on the y-axis. Light blue squares represent male samples, dark red circles represent female samples. The estimated intervention effect is plotted on the x-axis. Brackets indicate the 95% confidence interval. The p-values were adjusted for multiple testing using the false discovery rate. Significance is indicated by the asterisks (* = p.adjust < 0.05; ** = p.adjust < 0.01, *** = p.adjust < 0.001). Abbreviations: SAT; subcutaneous adipose tissue.

In general, we noted a similar behavior of the male/female eigengenes across the tissues (Figure 3). Remarkably, the eigengene of SAT-downregulated DEGs was upregulated in muscle (in both male and female) and the eigengene of muscle-upregulated DEGs was downregulated in SAT (although not significantly). Furthermore, the effect of the muscle and SAT eigengenes of DEGs were not significantly changed in blood, suggesting that these SAT/muscle genes were not affected by the intervention in blood. In summary, when investigating the intervention effect of the genes strongest affected by the intervention, no overlap of transcriptional response was observed between blood and SAT or muscle. Relationships did exist within the transcriptional response between SAT and muscle, although not always consistent in direction of effect.

### Especially SAT- and muscle-specific DEGs associate with health markers

To better understand the differential expressed transcriptome, we calculated the association between the eigengene values and the change in ten clinical metabolic health markers: *I*) two DEXA scan body composition measurements: whole-body fat%, trunk fat%; *II*) seven traditional metabolic health markers: BMI, Waist Circumference (WC) fasting insulin, fasting HDL cholesterol, fasting HDL cholesterol size, SBP, DBP; and *III*) the Framingham Risk Score (FRS), which is a cardiovascular risk score (Figure 4). Since the intervention effects on some of the health markers differed between the subgroups (Table 1), which thus can cause differences in effects due to differences in subsets, the relation between the expression data and metabolic health markers was performed on the overlapping blood, SAT and muscle samples (57; 33 males, 24 females). In blood, there were no significant associations between any of the eigengenes and classical metabolic health markers. The SAT eigengenes were significantly associated to the change in DEXA body composition parameters (whole-body fat%, trunk fat%), four of the seven traditional metabolic health markers (BMI, WC and fasting HDL cholesterol size) in the SAT transcriptome of both sexes, and with the fasting HDL cholesterol level and FRS only in men. Interestingly, also the muscle eigengenes expressed in SAT were significantly associated to the change in all of these parameters in men, except for fasting HDL cholesterol level, with the downregulated muscle eigengenes showing the strongest associations overall. In the transcriptome of muscle, the downregulated SAT eigengene associated negatively to DBP and the downregulated muscle eigengene associated to the change in DEXA body composition markers; whole-body fat%, trunk fat%, and metabolic health markers; BMI in men, BMI and WC in women.

**Figure 4:**
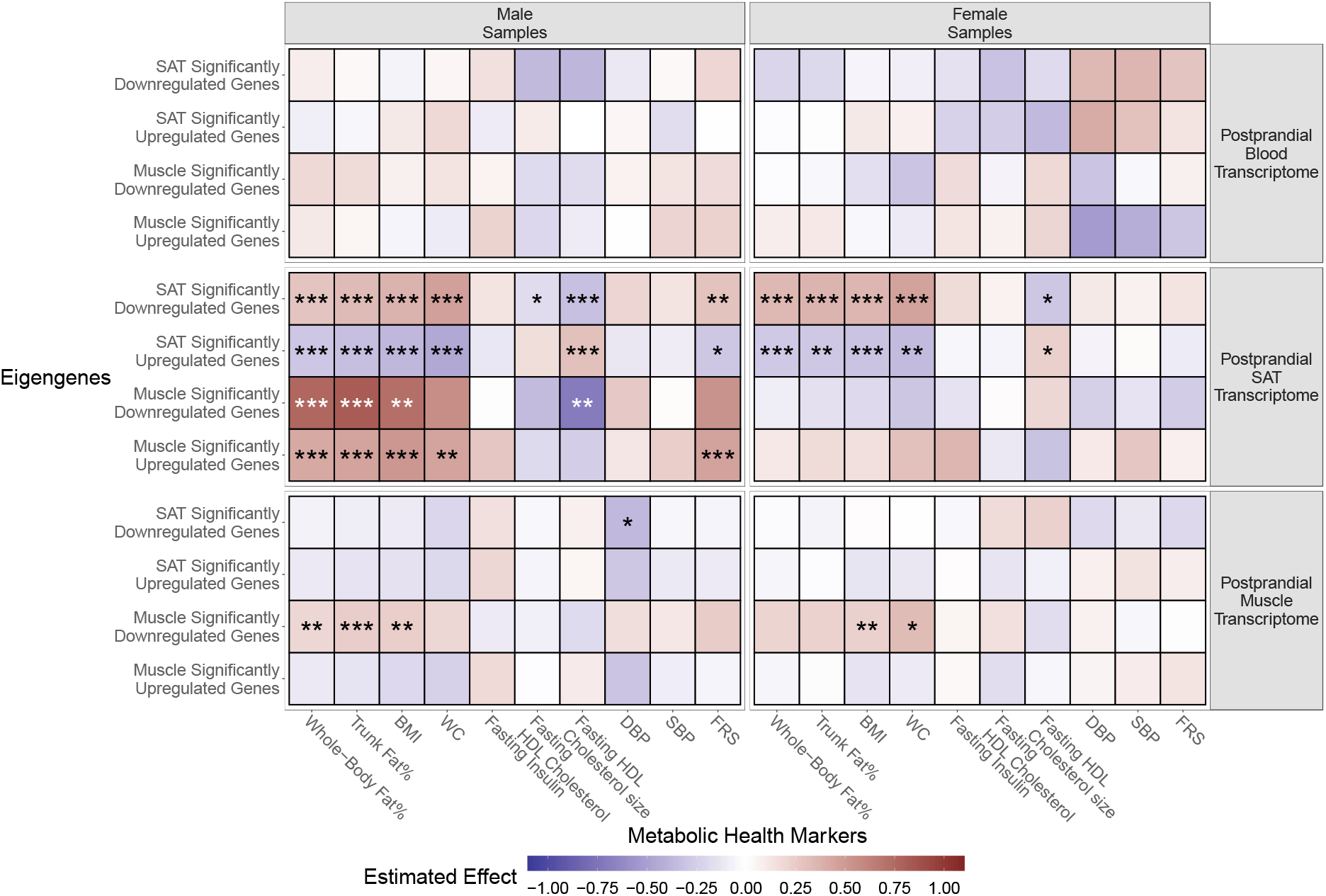
The SAT postprandial transcriptome captures the metabolic health changes stronger than the blood and muscle transcriptome. Genes used for input of the eigengenes were plotted on the y-axis. The metabolic health markers were plotted on the x-axis. Colors indicate the direction of the effect: blue, a negative effect, white; no effect, red; a positive effect, the darker the color, the stronger the effect. The p-values were adjusted for multiple testing using the false discovery rate. Significance is indicated by the asterisks (* = p.adjust < 0.05; ** = p.adjust < 0.01, *** = p.adjust < 0.001). Abbreviations: SAT; subcutaneous adipose tissue.

Taken together, especially in the SAT transcriptome, the SAT and muscle eigengenes reflected changes in DEXA body composition markers (whole-body and trunk fat%) and classical metabolic health markers (BMI, WC, fasting HDL cholesterol level and size), and to the former four also did the downregulated muscle eigengene in the muscle transcriptome. Many effects were similar in males and females, although more pronounced in males.

### Integrated analysis reveals male postprandial blood, SAT and muscle transcriptome share a joint variation

To explore the tissue dependency in a more intricate way, we captured the joint transcriptional reaction to the intervention across the three different tissues using Joint and Individual Variation Explained (JIVE) analysis (Materials and Methods, Supplementary Figure 1). The joint representation is a linear subspace spanned by the genes, analogous to a principal component analysis (PCA), but that now also explains the joint variation in expression in all three tissues across the intervention (Materials and Methods). Hence, although informed by all three tissues, the blood expression of a participant (before and after the intervention) can be mapped to this joint subspace without needing information about the muscle or SAT transcriptome. In other words, the blood transcriptome of a participant is decomposed in three (independent) transcriptome terms: 1) a term that reflects the joint transcriptomic variation in blood, SAT and muscle, 2) a term reflecting the remaining structured variation in blood only (also referred to as the individual variation), and 3) a term that represents the residual noise in the blood transcriptome, i.e. the signal that is not explained by the joint and individual terms. Subsequently, we can measure the differential expression across the intervention in either the joint subspace (of blood, SAT and muscle) or the individual subspaces (for each tissue types). The number of joint and individual components are determined through permutation testing (Materials and Methods).

In the male samples, JIVE identified 2 joint components for the shared transcriptomic variation in blood, SAT and muscle, and 11, 15 and 15 individual components for the blood, SAT and muscle, respectively (Supplementary Table 8). In postprandial blood and SAT, the joint components together explained 8% and 7.2% of the total variation in those tissues, respectively. In the postprandial muscle transcriptome, both joint components explained 31.3% of the variation. In female samples, JIVE did not identify a joint term that was independent from the individual terms (Supplementary Figure 2).

### In male samples, the drivers of the joint components were involved in phospholipid metabolism and the immune system

To investigate what genes expressed in postprandial blood were the drivers of the joint effect, the 250 genes that had the largest loading factors for each of the two joint components were selected, respectively (Materials and Methods). Both sets of genes were subsequently used in a pathway enrichment analysis. In postprandial blood, the genes driving the first joint component (JC1) that explains most of the joint variance across the tissues were involved phospholipid metabolism, while the genes driving the second joint component (JC2) were involved in interferon signaling, inflammation and other aspects of the immune system (Figure 5).

**Figure 5:**
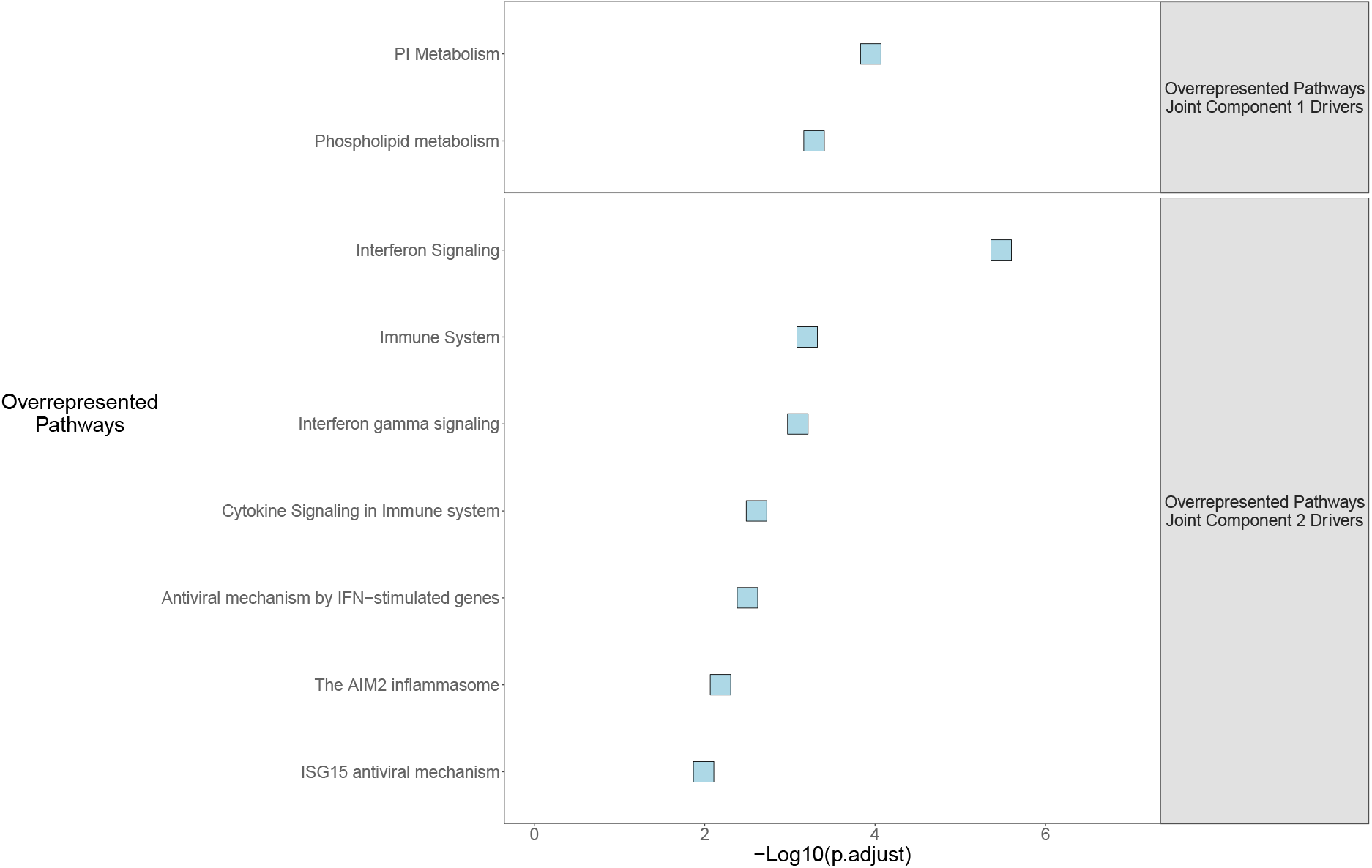
Significantly Overrepresented pathways in the top 250 drivers of the postprandial blood joint effect. The overrepresented pathways are plotted on the y-axis. Y-axis is ordered by significance, strongest significantly overrepresenting pathways first. The x-axis represents -log10 of the Bonferroni adjusted significance p-value.

### Phospholipid metabolism genes that drive the joint effect also capture the intervention effect individually

Next we wondered whether the genes driving the significant effects of JC1 and JC2 also showed an intervention effect by themselves. Within the both sets of 250 genes that had the highest loading factors for each of the joint components, we selected those genes that overlapped with the found enriched pathways. Interestingly, we found a remarkable difference between the intervention effect of the genes involved in phospholipid metabolism and those involved in immunity (Figure 6): the joint effect of the genes involved in phospholipid metabolism was more strongly influenced by the intervention than the individual effect, however, neither effect crossed the significant threshold after adjustment for multiple testing (Figure 6). Compared to the phospholipid metabolism genes, the effect sizes in both the joint nor individual effect of genes involved in the immune system were considerably weaker. These effects were also non-significantly associated with the intervention effect.

**Figure 6:**
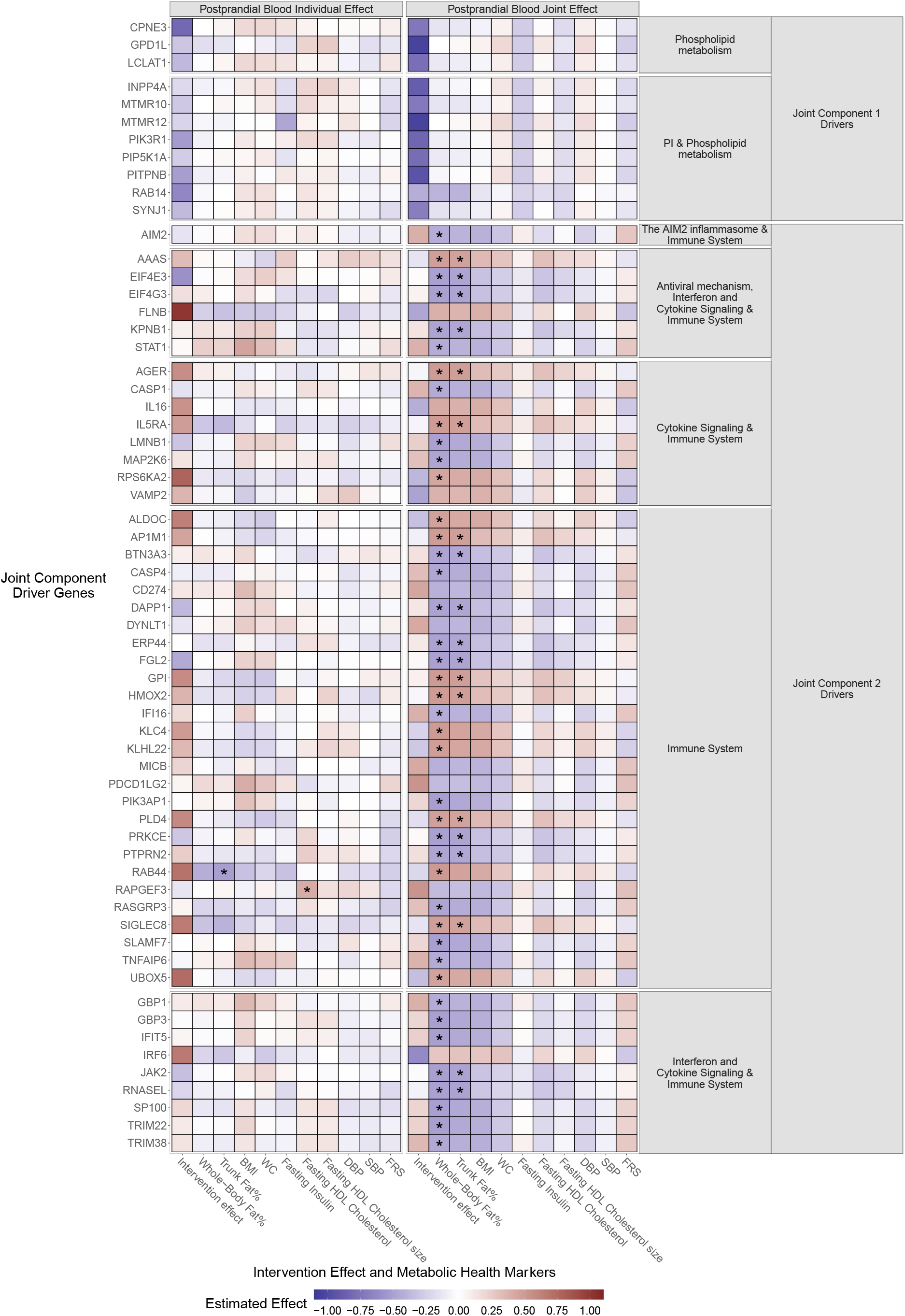
In postprandial blood, the joint component 1 drivers strongly captured the intervention effect while joint component 2 drivers was significantly associated to whole-body fat% and trunk fat% levels. The genes were plotted on the y-axis, the intervention effect and metabolic health markers were plotted on the x-axis. Colors indicate the direction of the effect: blue, a negative effect, white; no effect, red; a positive effect, the darker the color, the stronger the effect. The p-values were adjusted using the false discovery rate. Significance is indicated by the asterisks (* = p.adjust < 0.05).

### Immune genes that relate to the joint effect capture metabolic health effects of the intervention

Next we investigated whether changes in the genes involved in phospholipid metabolism and immunity associated with changes in the metabolic health markers. Here we found that neither the joint nor individual effect of the phospholipid genes associated to these health marker effects (Figure 6). In contrast, the joint effect of the genes involved in the immune system did strongly associate with markers of body composition (whole-body and trunk fat%) while, except for RAB44 and RAPGEF3 (which was associated to the fasting HDL cholesterol level), the individual effect did not. Hence, it is possible to detect effects of the intervention in the postprandial blood transcriptome, but only when the individual blood effect has been removed.

### Variance in the muscle and SAT transcriptome explain the selection of contributing genes to the joint components of blood differently

To investigate which of the two tissues, SAT or muscle, correlated most with the joint components in blood, we calculated the correlations of the top 250 genes with the highest loading factors for each of the joint components (JC1 and JC2) in blood with those in muscle and SAT (Supplementary Figure 3). The JC1 blood genes showed increasingly higher absolute correlations with the JC1 and JC2 muscle genes, whereas the JC2 blood genes showed an opposite behavior and were merely not correlated with the JC1 or JC2 muscle genes. On the other hand, the JC1 and JC2 SAT genes showed a more strong absolute correlation with the JC2 blood genes as opposed to the JC1 blood genes. These results indicate that the JC1 blood genes (dominated by genes involved in phospholipid metabolism) are largely correlated with the variance in the muscle transcriptome, the JC2 blood genes (dominated by genes involved in the immune system) are more correlated with the variances in the SAT transcriptome.

## Discussion

We investigated the effects in men and women of a mild combined lifestyle intervention in older adults on the postprandial transcriptome of blood, subcutaneous adipose tissue (SAT) and muscle tissue, and related the response to changes in parameters of metabolic health. Furthermore, using JIVE, we explored the shared variation between the three transcriptomes. In both sexes, the 13-week combined lifestyle intervention had virtually no effect on the postprandial blood transcriptome, while the postprandial SAT and muscle transcriptomes responded significantly. In postprandial SAT, pathways involved in HDL remodeling, cell signaling and CO2/O2 exchange (both sexes), ligand binding by scavenger receptors, NR1H3/NR1H2 cholesterol related gene expression and fertilization (male only), and signal transduction and MAPK signaling cascades (female only) were overrepresented, while in the postprandial muscle, pathways involved in collagen, extracellular matrix remodeling, protein O-glycolysation (male only) and osteoblast differentiation (female only) were significantly overexpressed.

The response of the SAT transcriptome in both sexes associated most strongly with changes in the Framing Risk Score (FRS), and metabolic health, including HDL cholesterol level and size, and specifically in body composition markers (whole-body and trunk fat%, BMI and Waist Circumference (WC)), and so did a part of the changes in the muscle transcriptome (whole-body and trunk fat%, BMI, WC and diastolic blood pressure (DBP)). We did observe sex differences in the association effect sizes. Lastly, we identified shared variation between the three transcriptomes, but only in the tissues of men. We identified in the blood transcriptome a joint intervention effect dominated by genes involved in phospholipid metabolism, largely influenced by genes overlapping with changes in the muscle transcriptome, and a second joint intervention effect dominated by genes involved in the immune system, overlapping with changes in the SAT transcriptome. The latter associated with changes in body composition (whole-body and trunk fat%). These effects were not detected in the original transcriptome of postprandial blood.

The effect of lifestyle interventions on the human transcriptome has mainly been reported for dietary and physical activity regimes separately, typically in metabolically unhealthy or frail participants ^22,24–26,30^. This makes a direct comparison with the response of the healthy, non-diabetic, older adults ^6^ to the GOTO combined lifestyle intervention difficult. Nevertheless, we do find similarities to dietary interventions. Like in GOTO, nutritional interventions with a mild energy restriction (20-25%, for 4-12 weeks) hardly affected the blood transcriptome ^24^. In adipose tissue, caloric restriction interventions changed the expression of genes involved in glucose and lipid metabolism, signal transduction, stem cell maintenance and vascularization ^18,21,25,26,30^, the former three of which were also overrepresented in the SAT transcriptome of the GOTO study (Figure 2). Lastly, in overweight or obese participants of a weight-loss intervention study (6 months, diet, physical activity separate and combined), genes involved in signaling pathways, translation, cell life span and ATP binding were significantly overrepresented ^31^, part of which we also identified in GOTO.

The effects of the muscle transcriptome to physical activity changes were reported in both anabolic and catabolic interventions, the latter being more similar to the regimen in GOTO. In line with the results found in GOTO, a systematic review investigating the effects of 21 activity interventions on the ECM in muscle, found that both types of activity interventions had a positive effect on collagen level and identified ECM adaptations related to muscle remodeling in both young and older participants ^27^. Not all gene expression effects are common to all interventions. A 6-months aerobic, resistance, or aerobic + resistance training regimen in obese older adults changed expression of genes involved in myogenesis, autophagy, mitochondrial biogenesis and inflammation, in all three training regimens ^23^, which was not observed for the muscle transcriptome in the GOTO study. This difference may be ascribed to the better health at baseline of GOTO individuals and/or the duration and intensity of this in essence anabolic intervention. Another anabolic 6-month resistance training intervention in frail and healthy older adults, showed effects similar to GOTO; an increase in expression levels of collagen and extracellular matrix related genes ^22^. Finally, in mice a combined caloric restriction and physical activity intervention reported DEGs involved in MAPK signaling, metabolic processes, cholesterol metabolism, extracellular matrix organization ^36^, which we also found in the GOTO study (Figure 2).

There is a number of other differences in study designs as compared to reported interventions. GOTO was a personalized intervention, to mimic a realistic situation when people change their lifestyle. Consequently, there was a considerable variation in the instructed lifestyle change between the participants; some changed their diet more, while other focused more in a change in physical activity. Furthermore the transcriptome in GOTO was measured in tissues taken after a standardized meal ^17^, which is usually not done.

A direct association between the expression levels in adipose tissue and muscle, and classical metabolic health markers is generally not looked into, making it difficult to relate the effects identified in GOTO (Figure 4). However, one cohort study of BMI discordant monozygotic twins (ΔBMI ≥ 2.5 kg/m2) reported on adipose tissue and skeletal muscle transcriptome differences ^41^. This study identified DEGs involved in ECM remodeling, DNA/RNA metabolism, mitochondrial metabolism (both adipose tissue and skeletal muscle), fatty acid metabolism, MAPK and AMPK signaling (adipose tissue only), and amino acid metabolism (skeletal muscle only) ^41^, which largely overlap with the overrepresented pathways in GOTO (Figure 2). Additionally, it was emphasized that combined differential gene expression scores of the adipose tissue transcriptome were stronger associated with classical metabolic health markers (including BMI, whole-body fat%, HDL cholesterol) than those of muscle transcriptome ^41^, which we also observed in GOTO (Figure 4).

When investigating the effects of the DEGs of one tissue in the other two tissues, we identified that the intervention had an effect on the SAT DEGs in the muscle transcriptome, but not the other way around (Figure 3). This indicated that the muscle transcriptome captured some of the effects on the SAT transcriptome, possibly by a SAT component in the muscle tissue.

Using the JIVE data integration approach ^40^, we have identified a shared variation between the three tissues in the male samples represented by two joint components (Supplementary Figures 1 & 2, Supplementary Table 8). The genes that drove these two joint components were involved in phospholipid metabolism and the immune system, respectively (Figure 5). Like the eigengene effects in the postprandial muscle transcriptome (Figure 3 and 4), the genes driving the first joint component (JC1) in blood showed a strong intervention effect but did not capture metabolic health effects (Figure 6). Moreover, these genes were also more correlated to variances in the muscle transcriptome than the SAT transcriptome (Supplementary Figure 3). Conversely, similarly to the effects in postprandial SAT, the genes driving the second joint component (JC2) in blood strongly captured the metabolic health effects of the intervention but had a weaker association to the intervention itself. This was also confirmed by a higher correlation with variances in the SAT transcriptome than the muscle transcriptome of the genes belonging to this second joint blood component (Supplementary Figure 3). From these results, we postulated that JC1 in blood was largely driven by the muscle transcriptome, while JC2 in blood was primarily driven by the SAT transcriptome.

This study faced several limitations, which need to be considered when interpreting the results. Firstly the number of participants with transcriptomics measurements of postprandial blood (88; 45 males, 24 females), postprandial SAT (78; 38 males, 40 females) and postprandial muscle (82; 48 males, 34 females), is relatively low, compared to the complete number of participants of the GOTO study (153; 75 males, 78 females). The variation in the instructed lifestyle changes in combination with the low number of samples and slight differences at baseline (Supplementary Table 1), could also have led to a large variation in outcomes when comparing participants with postprandial blood, SAT and muscle transcriptome measurements (Table 1). To circumvent part of this variation, the effects between the transcriptome and metabolic health markers were only studied within the participants with transcriptomic measurements in postprandial blood, SAT and muscle (57; 33 males, 24 females). Secondly, the group of participants with transcriptomic measurements of all three tissues was also used for the JIVE analysis. In females, the JIVE model did not identify a shared variation, this could be because of the shared effects between the tissues were too weak, which was likely attenuated by the low number of participants (24). In males, the JIVE analysis did identify two joint components between the transcriptomes of the three tissues.

Overall, we were able to identify considerable effects of a 13-week moderate nutritional and physical activity intervention on the transcriptome of SAT and muscle of healthy older adults. We demonstrated that the changes in expression of genes in metabolism pathways in SAT, and collagen and muscle remodeling pathways in muscle associated with beneficial features of overall metabolic health. These health effects were best captured by the postprandial SAT transcriptome. We identified a joint space of three tissues in postprandial blood, which was able to capture some of the effects in the other two tissues, even though postprandial blood alone did not capture this. We propose that this method could be used to estimate part of the effect of an intervention on SAT and muscle transcriptomes, when only blood expression data is available.

## Materials and Methods

### Study Design

The Growing Old tOgether (GOTO) study was a 13-week lifestyle intervention study with 164 healthy older adult participants (83 males, 81 females, mean age 62.9 +- 5.7, mean BMI 26.9 +- 2.5). The participants underwent a 25% reduction in energy expenditure, equally divided over decreased nutrient intake and increased physical activity, as described in ^6^. Both before and after the intervention, the participants were subjected to a nutrient challenge, consisting of an overnight fast and an intake of standardized nutrient shake, as described in ^17^. Thirty minutes after the nutrient challenge, samples of postprandial whole blood, postprandial subcutaneous adipose tissue (SAT) (*subcutaneous abdominal fat pad*) and postprandial muscle (*vastus vascularis*) were taken. Out of 164 participants, 153 (75 men, 78 women) were compliant to the intervention according to thresholds described in ^12^. Out of 153 compliant participants, postprandial transcriptomics were measured in three tissues: 88 (45 males, 43 females) in blood, 78 (38 males, 40 females) in SAT, 82 (48 males, 34 females) in muscle. Fifty-seven participants (33 males, 24 females) had postprandial transcriptomic measurements in all three tissues.

### Diagnostic Measurements

All measurements were performed in fasting serum collected through venipuncture. Insulin was measured using a Immulite 2000 xPi (Siemens, Eschborn, Germany). HOMA2-IR was calculated using the publicly available HOMA calculator (https://www.dtu.ox.ac.uk/homacalculator/) ^42^. Fasting HDL cholesterol and HDL cholesterol size were measured using the previously described Nightingale platform ^43^. Complete methods of diagnostic measurements are described in ^6^.

### DEXA measurements

Method for the DEXA measurements is described in ^14^. In short: whole-body DEXA (Discovery A, Hologic Inc., Bedford, MA,USA) was used to measure eleven body composition components, including whole-body fat mass and trunk fat mass. Whole-body fat% was calculated by dividing whole-body fat mass by whole-body mass. Trunk fat% was calculated by dividing the trunk fat mass by the total trunk weight.

### Investigation of Health Marker Effects

The intervention effect of each health marker was calculated using a linear mixed model, adjusted for age (fixed effects) and individual (random effects). The effect for males and females were calculated separately. The significance was adjusted for multiple testing using the Bonferroni correction method.

### RNA isolation and Sequencing

The method for RNA isolation and sequencing is described in ^17^. In short: libraries were prepared using Illumina TruSeq version 2 library preparation kits. Data processing was performed the in-house BIOPET Gentrap pipeline, described in ^44^. The following steps were part of the data processing: low quality trimming using sickle version 12.00. Cutadapt version 1.1 was used to perform the adapter clipping. The reads were aligned to GRCh37, while masking for SNPs common in the Dutch population (GoNL ^45^ MAF > 0.01), using STAR version 2.3.0e. Picard version 2.4.1. was used to perform sam to bam conversion and sorting. Read quantification was performed using htseq-count version 0.6.1.p1 using Ensemble gene annotations version 86 for gene definitions. In postprandial blood, the sequencing resulted in an average of 37.2 million reads per sample, 97% (+- 0.4%) of which were mapped. In postprandial SAT, samples had an average of 11.4 million sequenced reads, 95% (+- 1.6%) of which were mapped. In postprandial muscle, an average of 36.9 million sequence reads per sample, 98% (+- 0.4%) of which were mapped.

### Differential Gene Expression

In all tissues, the normalizations and differential gene expression analysis were performed in a sex stratified manner. Genes that had less than 2 counts on average, were removed from the dataset. Tissue specific confounders were identified by associationing them with the principal components that represent 90% of variation of TMM-CPM normalized gene expression.

#### Postprandial Blood

There were 88 participants (45 male, 43 females) with RNA-seq measurements pre -and post intervention. We adjusted for the following confounders: 1) technical effects: median 5’ bias, median 3’ bias, and flowcell, mean insert size in female samples only, 2) cell type percentages: eosinophiles, monocytes, lymphocytes, basophiles), and 3) personal details (age at baseline, and person ID as a random effect).

#### Postprandial SAT

78 participants (38 male, 40 female) had RNA-seq measurements pre-and post-intervention. We adjusted for the following confounders: 1) technical effects: molarity of sample, isolation date, and plate number in female samples only; and 2) personal measures: age at baseline, and person ID as a random effect.

#### Postprandial Muscle

There were 82 participants (48 male, 34 female) with RNA-seq measurements pre- and post-intervention. In male samples we adjusted for the following confouders: 1) technical effects: total yield, flow cell, isolations series, 28S/18S ratio, plate number, mean insert size, total number of aligned bases, RNA integrity number, median 5’ bias, median 3’ bias; and 2) personal details: age at baseline, and person ID as a random effect. In female samples, we adjusted for the following confounders: 1) technical effects: RNA 260/280 ratio, 28S/18S ratio, biopsy number, hand grip strength, RNA integrity number, mean insert size, isolation series, sequence % that passed quality control, total yield, total number of aligned bases, median 3’ bias; and 2) personal details: age at baseline, and person ID as a random effect.

The differential gene expression analysis was performed using linear mixed models analysis (limma version 3.52.2) in combination with VOOM normalization ^46^. We adjusted for multiple testing using the false discovery rate. An fdr-adjusted p-value < 0.01 was considered significant. Genes with an absolute log2 fold change ≥ 0.25 were considered differentially expressed.

### Functional Enrichment

The pathway overrepresentation analysis was performed using the enricher function of clusterProfiler (version 4.4.4) ^47^. Up-and downregulated significant DEGs were used as an input, all expressed genes that passed the quality control were used as a background. The Reactome database (version 81) ^48^ was used for the functional enrichment. The tests were adjusted for multiple testing using the Bonferroni correction method. An adjusted p-value below 0.05 was considered significant.

### Eigengene Analysis

The input for the eigengene calculation were the up- or downregulated significant DEGs. The eigengenes were calculated using the *prcomp* function of R package *stats* (version 4.2.1) ^49^, the first principal component was considered the eigengene. Eigengene values were scaled to mean 0 and standard deviation (sd) 1. The associations between the intervention (0 for baseline, 1 for post intervention) and the eigengenes was calculated with logistic regression using the function *glmer* of package *glmnet* (version 4.1-6) ^50^, adjusting for the same covariates as in the differential gene expression analysis. The significance level of the association was adjusted for multiple testing, using the false discovery rate. Similarly, the association between eigengenes and metabolic health markers are calculated. Metabolic health marker measurements were scaled to mean 0 and sd 1. Outliers more than 3 sd values from the mean were removed. Only participants with transcriptome measurements of all three tissues were used in the health marker association analysis.

### Joint and Individual Variation Explained (JIVE) Analysis

The Joint and Individual Variation Explained (JIVE) was calculated using the R package *r.jive* (version 2.4) ^40,51^. Analysis was performed on all genes that passed the quality control across 57 participants (33 Male, 24 Female). Prior to JIVE, the gene expression levels were scaled to a mean of 0 and a sd of 1. For each tissue matrix *X*^*t*^ ∈ ℝ^*p*^_*t*_ ^*x n*^ (where *t* represents the tissue, *p*_*t*_ represents the number of genes measured for tissue *t*, and *n* represents the number of samples), the JIVE analysis linearly decomposes the input matrix (*X*^*t*^) into three matrices representing: 1) the shared data across the tissue: *J*^*t*^ ∈ ℝ^*p*^_*t*_ ^*x n*^ explaining the joint variation across the three tissue types, 2) a data matrix representing the variation unique to a tissue: *A*^*t*^ ∈ ℝ^*p*^_*t*_ ^*x n*^, and 3) a data matrix representing the residual noise, i.e the remaining unexplained variation in a tissue: *R*^*t*^ ∈ ℝ^*p*^_*t*_ ^*x n*^, with: *X*^*t*^ = *J*^*t*^ + *A*^*t*^ + *R*^*t*^ (Supplementary Figure 1). *J*^*t*^ is further decomposed into sub-matrices *J*^*t*^ = *U*^*t*^ *S*, where *U*^*t*^ ∈ ℝ^*p*^_*t*_ ^*x r*^ represents a matrix containing the *p*_*t*_ gene loading vectors for the *r* joint components, and *S* is a sample scoring matrix of joint effects across all tissues (*S* is the same for all tissues). The number of joint components, *r*, is selected using permutation testing as described in ^40^. Likewise, *A*^*t*^is decomposed, and for each tissue, *A*^*t*^ = *W*^*t*^ *S*^*t*^, where *W*^*t*^ ∈ ℝ^*p*^_*t*_ ^*x rt*^ represents a matrix containing the *p*_*t*_ gene loading vectors for the *r*_*t*_ individual tissue components (selected by permutation testing), and *S*^*t*^ ∈ ℝ^*r*^_*t*_ ^*x n*^ is a sample scoring matrix of individual components. For each tissue, we can identify the genes that contribute most to the joint and individual effects based on the ranking of genes by each column of *U*^*t*^and *W*^*t*^, respectively.

## Data Availability Statement

The data that support the findings of this study are accessible upon request to the corresponding author. The data are not publicly available due to privacy or ethical restrictions.

## Author Contributions

P.E. Slagboom and C.P.G.M. de Groot designed the study. M. Beekman collected and curated the health data. N. Lakenberg and E. Suchiman generated the transcriptome data. F.A. Bogaards T. Gehrmann, M.J.T. Reinders and P.E. Slagboom designed the data analysis approach, F.A. Bogaards, performed data analysis and F.A. Bogaards, T. Gehrmann, M.J.T. Reinders, C.P.G.M. de Groot and P.E. Slagboom performed the research and interpreted the data. All authors were involved in drafting and revising the manuscript.

## Ethics Statement

The Medical Ethical Committee of the Leiden University Medical Center approved the study (P11.187) and all participants signed a written informed consent. All experiments were performed in accordance with relevant and approved guidelines and regulations. This trial was registered at the Dutch Trial Register (http://www.trialregister.nl) as NTR3499.

## Funding and Acknowledgements

This work was funded by the Horizon 2020 ERC Advanced grant: GEROPROTECT, the Netherlands Consortium for Healthy Ageing (NWO grant 050-060-810), the framework of the BBMRI Metabolomics Consortium funded by BBMRI-NL (NWO 184.021.007 and 184.033.111), ZonMw Project VOILA (ZonMW 457001001) and X-omics (NWO 184.034.019). The funding agencies had no role in the design and conduct of the study; collection, management, analysis, and interpretation of the data; and preparation, review, or approval of the manuscript. The authors would like to express their gratitude to all participants of the GOTO study who did their very best to adhere to the intervention guidelines and underwent all measurements.

