## Supplements for "Transcriptomic Response of Postprandial Blood, Subcutaneous Adipose Tissue and Muscle to a combined lifestyle intervention in older adults"

Supplementary Information

***Supplementary Table 1: Baseline measurements of metabolic health marker measurements per subset selection.***

| Health Marker, baseline mean (SD) | Sex | Complete GOTO dataset 153 (75 Male, 78 Female) | Overlapping Blood, SAT and Muscle Samples 57 (33 Male, 24 Female) | Blood Postprandial Samples 88 (45 Male, 43 Female) | SAT Postprandial Samples 78 (38 Male, 40 Female) | Muscle Postprandial Samples 82 (48 Male, 34 Female) |
|---|---|---|---|---|---|---|
| Age, years | Male | 63.95 (5.36) | 65.08 (5.65) | 64.21 (5.68) | 64.86 (5.51) | 64.4 (5.5) |
| BMI, kg/m$^2$ | Male | 26.53 (1.95) | 26.58 (2.04) | 26.59 (2) | 26.78 (2.19) | 26.6 (2.05) |
| Waist Circumference | Male | 98.2 (6.47) | 99.27 (6.03) | 98.6 (6.39) | 99.58 (6.09) | 98.9 (6.56) |
| Systolic Blood Pressure, mm Hg | Male | 86.22 (7.52) | 87.66 (7.99) | 86.87 (7.58) | 87.74 (7.91) | 86.79 (7.64) |
| Diastolic Blood Pressure, mm Hg | Male | 141.02 (14.84) | 142.95 (14.67) | 140.46 (15.13) | 142.36 (14.44) | 140.95 (14.76) |
| Fasting Insulin, mU/L | Male | 9.42 (4.06) | 9.39 (3.23) | 9.07 (3.24) | 9.69 (3.57) | 9.36 (3.59) |
| Whole-Body Fat, % | Male | 26.5 (4.37) | 26.76 (4.64) | 26.25 (4.7) | 27.02 (4.71) | 26.5 (4.77) |
| Trunk Fat, % | Male | 28.17 (5.38) | 28.09 (5.33) | 27.55 (5.68) | 28.42 (5.56) | 27.85 (5.84) |
| Fasting HDL Cholesterol, mmol/L | Male | 1.36 (0.23) | 1.38 (0.24) | 1.36 (0.24) | 1.38 (0.23) | 1.36 (0.24) |
| Fasting HDL Cholesterol size, nm | Male | 9.49 (0.14) | 9.49 (0.15) | 9.49 (0.14) | 9.5 (0.14) | 9.49 (0.14) |
| FRS | Male | 12.96 (1.54) | 13.21 (1.29) | 12.98 (1.44) | 13.11 (1.35) | 13.04 (1.35) |
| Age, years | Female | 62.02 (6) | 61.83 (4.84) | 62.74 (5.4) | 61.86 (5.27) | 61.99 (4.77) |
| BMI, kg/m$^2$ | Female | 27.04 (2.73) | 26.86 (2.68) | 26.67 (2.34) | 27.21 (2.68) | 26.85 (2.66) |
| Waist Circumference | Female | 93.36 (7.83) | 93.33 (7.2) | 93.14 (7.48) | 95.1 (7.28) | 93.68 (7.35) |
| Systolic Blood Pressure, mm Hg | Female | 84.23 (8.6) | 85.18 (9.14) | 84.07 (8.46) | 85.89 (8.47) | 84.66 (9.23) |
| Diastolic Blood Pressure, mm Hg | Female | 137.72 (17.27) | 138.16 (17.06) | 136.05 (16.37) | 137.59 (15.84) | 136.48 (15.96) |
| Fasting Insulin, mU/L | Female | 8.35 (3.99) | 7.78 (3.2) | 7.57 (3.59) | 8.24 (4.12) | 7.69 (3.25) |
| Whole-Body Fat, % | Female | 38.16 (4.39) | 37.34 (4.81) | 37.7 (4.24) | 37.85 (4.46) | 37.19 (4.92) |
| Trunk Fat, % | Female | 37.4 (5.55) | 36.93 (5.82) | 36.82 (5.18) | 37.3 (5.79) | 36.72 (6.2) |
| Fasting HDL Cholesterol, mmol/L | Female | 1.64 (0.28) | 1.64 (0.27) | 1.68 (0.29) | 1.66 (0.26) | 1.64 (0.26) |
| Fasting HDL Cholesterol size, nm | Female | 9.62 (0.17) | 9.59 (0.16) | 9.62 (0.17) | 9.61 (0.16) | 9.62 (0.17) |
| FRS | Female | 14.88 (2.72) | 14.88 (2.58) | 14.91 (2.56) | 14.8 (2.33) | 14.71 (2.3) |

*Abbreviations: SD; standard deviation, SAT; subcutaneous adipose tissue, BMI; body mass index, FRS; Framingham Risk Score*

***Supplementary Table 2: Significantly differentially expressed genes in Male Postprandial Blood Samples***

| Ensembl ID | Gene Symbol | log2FC | FDR adjusted P-value |
|---|---|---|---|
| ENSG00000089163 | SIRT4 | -0.254671481895593 | 7.84E-03 |

***Supplementary Table 3: Significantly differentially expressed genes in Male Postprandial SAT Samples***

| Ensembl ID | Gene Symbol | log2FC | FDR adjusted P-value |
|---|---|---|---|
| ENSG00000263874 | LINC00672 | -0.5672204 | 1.32E-05 |
| ENSG00000124103 | FAM209A | -0.3910535 | 1.31E-04 |
| ENSG00000168994 | PXDC1 | -0.3382008 | 1.68E-04 |
| ENSG00000234072 | AC074117.10 | 0.26464397 | 2.59E-04 |
| ENSG00000135698 | MPHOSPH6 | 0.25210755 | 7.10E-04 |
| ENSG00000225828 | FAM229A | -0.2520179 | 7.10E-04 |
| ENSG00000143367 | TUFT1 | -0.3671268 | 7.10E-04 |
| ENSG00000101955 | SRPX | 0.34807962 | 7.10E-04 |
| ENSG00000144381 | HSPD1 | 0.26412275 | 7.10E-04 |
| ENSG00000183010 | PYCR1 | 0.49283832 | 7.54E-04 |
| ENSG00000130203 | APOE | 0.53982265 | 7.54E-04 |
| ENSG00000161048 | NAPEPLD | 0.25767271 | 7.54E-04 |
| ENSG00000115325 | DOK1 | -0.2515008 | 8.99E-04 |
| ENSG00000113140 | SPARC | -0.2718733 | 8.99E-04 |
| ENSG00000111087 | GLI1 | -0.400404 | 1.07E-03 |
| ENSG00000135441 | BLOC1S1 | -0.4035352 | 1.07E-03 |
| ENSG00000250303 | RP11-356J5.12 | -0.3754066 | 1.10E-03 |
| ENSG00000163884 | KLF15 | 0.38622015 | 1.10E-03 |
| ENSG00000000005 | TNMD | -0.4330151 | 1.39E-03 |
| ENSG00000165804 | ZNF219 | -0.283069 | 1.74E-03 |
| ENSG00000261888 | AC144831.1 | -0.4213736 | 1.89E-03 |
| ENSG00000167566 | NCKAP5L | -0.2935398 | 2.16E-03 |
| ENSG00000233688 | CTD-2183H9.3 | 0.39407562 | 2.21E-03 |
| ENSG00000244734 | HBB | -1.2647543 | 2.21E-03 |
| ENSG00000130208 | APOC1 | 0.6870979 | 2.28E-03 |
| ENSG00000065054 | SLC9A3R2 | 0.34183357 | 2.28E-03 |
| ENSG00000164082 | GRM2 | -0.4737152 | 2.28E-03 |
| ENSG00000260105 | AOC4P | -0.4268106 | 2.41E-03 |
| ENSG00000137124 | ALDH1B1 | 0.25343564 | 2.41E-03 |
| ENSG00000100312 | ACR | -0.5064562 | 2.41E-03 |
| ENSG00000260025 | RP11-490M8.1 | -0.2781459 | 2.50E-03 |
| ENSG00000163638 | ADAMTS9 | 0.33946403 | 2.84E-03 |
| ENSG00000224003 | YES1P1 | 0.39589006 | 2.93E-03 |
| ENSG00000128965 | CHAC1 | -0.7184311 | 2.93E-03 |
| ENSG00000012171 | SEMA3B | -0.4258316 | 3.08E-03 |

| | | | |
|---|---|---|---|
| ENSG00000168140 | VASN | -0.272428 | 3.27E-03 |
| ENSG00000184163 | FAM132A | -0.3552164 | 3.27E-03 |
| ENSG00000164035 | EMCN | 0.25803273 | 3.51E-03 |
| ENSG00000259895 | RP11-715J22.2 | 0.36282949 | 3.51E-03 |
| ENSG00000099957 | P2RX6 | -0.5127287 | 3.51E-03 |
| ENSG00000164112 | TMEM155 | -0.3838115 | 3.56E-03 |
| ENSG00000251023 | RP11-549J18.1 | 0.27293591 | 3.60E-03 |
| ENSG00000143171 | RXRG | 0.36394195 | 4.09E-03 |
| ENSG00000143845 | ETNK2 | -0.3765187 | 4.12E-03 |
| ENSG00000102755 | FLT1 | 0.27988016 | 4.23E-03 |
| ENSG00000108179 | PPIF | 0.38363987 | 4.27E-03 |
| ENSG00000188536 | HBA2 | -1.2597373 | 4.27E-03 |
| ENSG00000165917 | RAPSN | -0.3768796 | 4.43E-03 |
| ENSG00000146477 | SLC22A3 | 0.29401595 | 4.68E-03 |
| ENSG00000237781 | RP11-54A4.2 | -0.3727969 | 4.70E-03 |
| ENSG00000003249 | DBNDD1 | -0.4208892 | 4.70E-03 |
| ENSG00000132688 | NES | 0.35652936 | 4.96E-03 |
| ENSG00000145861 | C1QTNF2 | -0.3581016 | 4.98E-03 |
| ENSG00000183615 | FAM167B | 0.3176645 | 5.14E-03 |
| ENSG00000232324 | AC008440.10 | -0.3299187 | 5.19E-03 |
| ENSG00000158578 | ALAS2 | -0.5996986 | 5.29E-03 |
| ENSG00000003137 | CYP26B1 | -0.2933832 | 5.29E-03 |
| ENSG00000115541 | HSPE1 | 0.29785867 | 5.54E-03 |
| ENSG00000268166 | NA | 0.28471935 | 5.54E-03 |
| ENSG00000087237 | CETP | 0.75391346 | 5.54E-03 |
| ENSG00000119681 | LTBP2 | -0.3288504 | 6.00E-03 |
| ENSG00000120254 | MTHFD1L | 0.27459313 | 6.01E-03 |
| ENSG00000171227 | TMEM37 | -0.2710702 | 6.09E-03 |
| ENSG00000188112 | C6orf132 | -0.3980761 | 6.13E-03 |
| ENSG00000111262 | KCNA1 | -0.3686729 | 6.65E-03 |
| ENSG00000120694 | HSPH1 | 0.26593407 | 6.65E-03 |
| ENSG00000020577 | SAMD4A | -0.2551815 | 6.65E-03 |
| ENSG00000270021 | CTC-203F4.2 | -0.2965268 | 6.65E-03 |
| ENSG00000137168 | PPIL1 | 0.26184583 | 6.71E-03 |
| ENSG00000004478 | FKBP4 | 0.2534673 | 6.73E-03 |
| ENSG00000129474 | AJUBA | -0.2739182 | 6.88E-03 |
| ENSG00000176402 | GJC3 | 0.40427611 | 6.97E-03 |
| ENSG00000267838 | AC008746.12 | 0.34251154 | 7.02E-03 |
| ENSG00000247033 | RP11-252E2.1 | 0.35716562 | 7.04E-03 |
| ENSG00000099338 | CATSPERG | -0.3461822 | 7.14E-03 |
| ENSG00000269837 | IPO5P1 | 0.2531476 | 7.41E-03 |

| | | | |
|---|---|---|---|
| ENSG00000267532 | MIR497HG | 0.36692844 | 7.41E-03 |
| ENSG00000149506 | ZP1 | 0.38184648 | 7.52E-03 |
| ENSG00000203392 | CTD-2026K11.6 | -0.3280197 | 7.64E-03 |
| ENSG00000174236 | REP15 | 0.29423172 | 7.87E-03 |
| ENSG00000176845 | METRNL | -0.2552482 | 7.98E-03 |
| ENSG00000227268 | KLLN | 0.33719879 | 8.44E-03 |
| ENSG00000264868 | NA | 0.26990175 | 8.50E-03 |
| ENSG00000105369 | CD79A | -0.4457534 | 8.60E-03 |
| ENSG00000166292 | TMEM100 | 0.28901649 | 8.65E-03 |
| ENSG00000148926 | ADM | -0.2650528 | 8.85E-03 |
| ENSG00000173641 | HSPB7 | -0.2738556 | 9.01E-03 |
| ENSG00000130518 | KIAA1683 | -0.3112072 | 9.32E-03 |
| ENSG00000258900 | HNRNPCP1 | 0.28994722 | 9.58E-03 |

***Supplementary Table 4: Significantly differentially expressed genes in Male Postprandial Muscle Samples***

| Ensembl ID | Gene Symbol | log2FC | FDR adjusted P-value |
|---|---|---|---|
| ENSG00000087303 | NID2 | 0.60428388 | 3.54E-04 |
| ENSG00000108821 | COL1A1 | 1.86558937 | 3.54E-04 |
| ENSG00000168542 | COL3A1 | 1.53342063 | 3.54E-04 |
| ENSG00000149257 | SERPINH1 | 0.72930014 | 3.54E-04 |
| ENSG00000087116 | ADAMTS2 | 1.33377375 | 3.54E-04 |
| ENSG00000134962 | KLB | -0.4235069 | 7.06E-04 |
| ENSG00000249825 | CTD-2201I18.1 | 0.66759779 | 7.07E-04 |
| ENSG00000197283 | SYNGAP1 | 0.41055465 | 8.17E-04 |
| ENSG00000139289 | PHLDA1 | 0.49719069 | 1.00E-03 |
| ENSG00000113296 | THBS4 | 0.88434736 | 1.33E-03 |
| ENSG00000088882 | CPXM1 | 1.27625498 | 1.33E-03 |
| ENSG00000077274 | CAPN6 | 0.95783851 | 1.33E-03 |
| ENSG00000204262 | COL5A2 | 1.15165247 | 1.33E-03 |
| ENSG00000166997 | CNPY4 | 0.38365822 | 1.33E-03 |
| ENSG00000162366 | PDZK1IP1 | -0.3535748 | 1.33E-03 |
| ENSG00000106484 | MEST | 0.89283927 | 1.33E-03 |
| ENSG00000164692 | COL1A2 | 1.13568329 | 1.33E-03 |
| ENSG00000168246 | UBTD2 | 0.26402582 | 1.33E-03 |
| ENSG00000242265 | PEG10 | 0.5091535 | 1.33E-03 |
| ENSG00000083828 | ZNF586 | -0.2909243 | 1.33E-03 |
| ENSG00000115129 | TP53I3 | 0.52097644 | 1.33E-03 |
| ENSG00000026652 | AGPAT4 | 0.40594296 | 1.33E-03 |
| ENSG00000106571 | GLI3 | 0.48943752 | 1.33E-03 |
| ENSG00000129749 | CHRNA10 | -0.2638822 | 1.33E-03 |
| ENSG00000142408 | CACNG8 | 0.35382717 | 1.33E-03 |
| ENSG00000130508 | PXDN | 0.58469698 | 1.33E-03 |
| ENSG00000171365 | CLCN5 | 0.42250447 | 1.33E-03 |
| ENSG00000187498 | COL4A1 | 0.62993361 | 1.33E-03 |
| ENSG00000121005 | CRISPLD1 | 0.586018 | 1.33E-03 |
| ENSG00000101825 | MXRA5 | 1.35661349 | 1.33E-03 |
| ENSG00000102316 | MAGED2 | 0.26899677 | 1.33E-03 |
| ENSG00000218891 | ZNF579 | -0.3298048 | 1.33E-03 |
| ENSG00000101955 | SRPX | 0.61148377 | 1.34E-03 |
| ENSG00000130635 | COL5A1 | 1.07808768 | 1.47E-03 |
| ENSG00000166963 | MAP1A | 0.62951012 | 1.47E-03 |
| ENSG00000134013 | LOXL2 | 0.83048007 | 1.61E-03 |
| ENSG00000173950 | XXYLT1 | 0.36125903 | 1.61E-03 |

| | | | |
|---|---|---|---|
| ENSG00000117385 | P3H1 | 0.41150445 | 1.76E-03 |
| ENSG00000188536 | HBA2 | -1.3454209 | 1.76E-03 |
| ENSG00000138463 | DIRC2 | -0.2507049 | 1.76E-03 |
| ENSG00000154096 | THY1 | 1.01203111 | 1.83E-03 |
| ENSG00000091986 | CCDC80 | 0.94939853 | 1.89E-03 |
| ENSG00000187955 | COL14A1 | 1.15312388 | 1.94E-03 |
| ENSG00000017427 | IGF1 | 0.69090611 | 1.94E-03 |
| ENSG00000134871 | COL4A2 | 0.57761461 | 1.96E-03 |
| ENSG00000162944 | RFTN2 | 0.31692045 | 2.06E-03 |
| ENSG00000174697 | LEP | -0.6471973 | 2.06E-03 |
| ENSG00000184897 | H1FX | -0.2587903 | 2.12E-03 |
| ENSG00000197093 | GAL3ST4 | 0.43689088 | 2.12E-03 |
| ENSG00000109436 | TBC1D9 | 0.30855514 | 2.20E-03 |
| ENSG00000117594 | HSD11B1 | -0.4782281 | 2.42E-03 |
| ENSG00000131747 | TOP2A | 0.39395638 | 2.52E-03 |
| ENSG00000171864 | PRND | 0.43051362 | 2.57E-03 |
| ENSG00000106819 | ASPN | 0.98080725 | 2.66E-03 |
| ENSG00000168916 | ZNF608 | 0.38464378 | 2.74E-03 |
| ENSG00000196730 | DAPK1 | 0.39366936 | 2.74E-03 |
| ENSG00000112769 | LAMA4 | 0.49486352 | 2.74E-03 |
| ENSG00000206384 | COL6A6 | 0.6839613 | 2.74E-03 |
| ENSG00000206172 | HBA1 | -0.8927129 | 2.74E-03 |
| ENSG00000127863 | TNFRSF19 | 0.50266853 | 2.74E-03 |
| ENSG00000146966 | DENND2A | 0.2825735 | 2.74E-03 |
| ENSG00000164099 | PRSS12 | 0.57471518 | 2.74E-03 |
| ENSG00000167123 | CERCAM | 0.69875573 | 3.01E-03 |
| ENSG00000082196 | C1QTNF3 | 0.594426 | 3.07E-03 |
| ENSG00000163359 | COL6A3 | 0.6990966 | 3.10E-03 |
| ENSG00000123643 | SLC36A1 | 0.39820748 | 3.10E-03 |
| ENSG00000135821 | GLUL | -0.410759 | 3.10E-03 |
| ENSG00000113083 | LOX | 0.69495176 | 3.12E-03 |
| ENSG00000184811 | TUSC5 | -0.6807371 | 3.20E-03 |
| ENSG00000049540 | ELN | 1.13845447 | 3.20E-03 |
| ENSG00000125089 | SH3TC1 | 0.3753011 | 3.20E-03 |
| ENSG00000155893 | PXYLP1 | 0.32511057 | 3.20E-03 |
| ENSG00000165029 | ABCA1 | 0.38084133 | 3.20E-03 |
| ENSG00000065308 | TRAM2 | 0.27507816 | 3.20E-03 |
| ENSG00000113140 | SPARC | 0.57926331 | 3.29E-03 |
| ENSG00000168890 | TMEM150A | -0.2529654 | 3.35E-03 |
| ENSG00000099875 | MKNK2 | -0.2611496 | 3.40E-03 |
| ENSG00000164237 | CMBL | -0.2898665 | 3.40E-03 |
| ENSG00000120278 | PLEKHG1 | 0.3003319 | 3.50E-03 |

| | | | |
|---|---|---|---|
| ENSG00000168939 | SPRY3 | 0.26110452 | 3.53E-03 |
| ENSG00000164294 | GPX8 | 0.54806961 | 3.54E-03 |
| ENSG00000164694 | FNDC1 | 1.06587158 | 3.54E-03 |
| ENSG00000084774 | CAD | 0.26264087 | 3.54E-03 |
| ENSG00000111602 | TIMELESS | 0.2515423 | 3.60E-03 |
| ENSG00000162745 | OLFML2B | 0.70961633 | 3.70E-03 |
| ENSG00000164946 | FREM1 | 0.43030329 | 3.81E-03 |
| ENSG00000105472 | CLEC11A | 0.36040905 | 3.81E-03 |
| ENSG00000157833 | GAREM2 | -0.3972877 | 3.81E-03 |
| ENSG00000204099 | NEU4 | 0.55747358 | 3.81E-03 |
| ENSG00000115252 | PDE1A | 0.36715315 | 3.94E-03 |
| ENSG00000149633 | KIAA1755 | 0.46449044 | 3.94E-03 |
| ENSG00000204291 | COL15A1 | 0.51648402 | 3.98E-03 |
| ENSG00000072832 | CRMP1 | 0.4056849 | 3.98E-03 |
| ENSG00000183386 | FHL3 | -0.3454115 | 4.01E-03 |
| ENSG00000184232 | OAF | 0.44462158 | 4.01E-03 |
| ENSG00000164116 | GUCY1A3 | 0.31716861 | 4.01E-03 |
| ENSG00000164125 | FAM198B | 0.43358407 | 4.01E-03 |
| ENSG00000230454 | U73166.2 | 0.26197833 | 4.01E-03 |
| ENSG00000072195 | SPEG | -0.2931974 | 4.01E-03 |
| ENSG00000140937 | CDH11 | 0.63957665 | 4.01E-03 |
| ENSG00000164855 | TMEM184A | -0.3103086 | 4.01E-03 |
| ENSG00000138801 | PAPSS1 | 0.32416866 | 4.01E-03 |
| ENSG00000088986 | DYNLL1 | 0.31712345 | 4.01E-03 |
| ENSG00000138119 | MYOF | 0.51878408 | 4.01E-03 |
| ENSG00000116962 | NID1 | 0.44809705 | 4.17E-03 |
| ENSG00000136378 | ADAMTS7 | 0.4974071 | 4.17E-03 |
| ENSG00000133138 | TBC1D8B | 0.30930514 | 4.17E-03 |
| ENSG00000114115 | RBP1 | 0.51836892 | 4.18E-03 |
| ENSG00000140986 | RPL3L | -0.2587998 | 4.21E-03 |
| ENSG00000163430 | FSTL1 | 0.63531771 | 4.29E-03 |
| ENSG00000178821 | TMEM52 | -0.3177182 | 4.29E-03 |
| ENSG00000187193 | MT1X | -0.4114528 | 4.29E-03 |
| ENSG00000133026 | MYH10 | 0.32356073 | 4.29E-03 |
| ENSG00000091136 | LAMB1 | 0.46762795 | 4.30E-03 |
| ENSG00000138678 | GPAT3 | -0.3082461 | 4.30E-03 |
| ENSG00000145423 | SFRP2 | 1.09628552 | 4.30E-03 |
| ENSG00000117600 | PLPPR4 | 0.58268307 | 4.30E-03 |
| ENSG00000169515 | CCDC8 | 0.49480992 | 4.32E-03 |
| ENSG00000145685 | LHFPL2 | 0.40610093 | 4.42E-03 |
| ENSG00000163565 | IFI16 | 0.41794575 | 4.45E-03 |
| ENSG00000174705 | SH3PXD2B | 0.62864955 | 4.46E-03 |

| | | | |
|---|---|---|---|
| ENSG00000142156 | COL6A1 | 0.62468728 | 4.48E-03 |
| ENSG00000196839 | ADA | 0.32848431 | 4.50E-03 |
| ENSG00000153885 | KCTD15 | 0.33270013 | 4.50E-03 |
| ENSG00000115155 | OTOF | 0.37605831 | 4.50E-03 |
| ENSG00000092969 | TGFB2 | 0.36873251 | 4.50E-03 |
| ENSG00000129744 | ART1 | -0.2512717 | 4.50E-03 |
| ENSG00000200201 | Y_RNA | -0.2648446 | 4.55E-03 |
| ENSG00000172296 | SPTLC3 | 0.25986582 | 4.55E-03 |
| ENSG00000184408 | KCND2 | 0.35099699 | 4.61E-03 |
| ENSG00000134247 | PTGFRN | 0.55462441 | 4.63E-03 |
| ENSG00000112541 | PDE10A | 0.32050209 | 4.63E-03 |
| ENSG00000165124 | SVEP1 | 0.47727976 | 4.65E-03 |
| ENSG00000178904 | DPY19L3 | 0.26922258 | 4.65E-03 |
| ENSG00000100362 | PVALB | 1.01993189 | 4.65E-03 |
| ENSG00000182326 | C1S | 0.44978186 | 4.69E-03 |
| ENSG00000010295 | IFFO1 | 0.28963872 | 4.69E-03 |
| ENSG00000219481 | NBPF1 | 0.3354468 | 4.69E-03 |
| ENSG00000146242 | TPBG | 0.34813091 | 4.69E-03 |
| ENSG00000139329 | LUM | 0.81727195 | 4.69E-03 |
| ENSG00000143320 | CRABP2 | 0.74169202 | 4.71E-03 |
| ENSG00000182492 | BGN | 0.80604944 | 4.82E-03 |
| ENSG00000204420 | C6orf25 | -0.2660635 | 4.83E-03 |
| ENSG00000183853 | KIRREL | 0.34177527 | 4.83E-03 |
| ENSG00000161381 | PLXDC1 | 0.41116466 | 4.85E-03 |
| ENSG00000143344 | RGL1 | 0.46719422 | 4.88E-03 |
| ENSG00000103855 | CD276 | 0.45212509 | 4.97E-03 |
| ENSG00000034510 | TMSB10 | 0.30721006 | 5.05E-03 |
| ENSG00000075618 | FSCN1 | 0.48708893 | 5.09E-03 |
| ENSG00000182463 | TSHZ2 | 0.37745064 | 5.09E-03 |
| ENSG00000211448 | DIO2 | 0.51763893 | 5.12E-03 |
| ENSG00000197496 | SLC2A10 | 0.44180945 | 5.12E-03 |
| ENSG00000170004 | CHD3 | 0.36436536 | 5.12E-03 |
| ENSG00000166033 | HTRA1 | 0.56614381 | 5.12E-03 |
| ENSG00000106809 | OGN | 0.94094545 | 5.12E-03 |
| ENSG00000162627 | SNX7 | 0.52129854 | 5.20E-03 |
| ENSG00000113328 | CCNG1 | -0.2801789 | 5.24E-03 |
| ENSG00000087245 | MMP2 | 0.67790037 | 5.28E-03 |
| ENSG00000006042 | TMEM98 | 0.45412643 | 5.28E-03 |
| ENSG00000126218 | F10 | 0.35167079 | 5.29E-03 |
| ENSG00000158270 | COLEC12 | 0.48724515 | 5.30E-03 |
| ENSG00000106483 | SFRP4 | 0.92280762 | 5.36E-03 |
| ENSG00000244734 | HBB | -1.2610097 | 5.57E-03 |

| | | | |
|---|---|---|---|
| ENSG00000106688 | SLC1A1 | -0.2676131 | 5.78E-03 |
| ENSG00000107859 | PITX3 | -0.3057413 | 5.78E-03 |
| ENSG00000121764 | HCRTR1 | -0.2604659 | 5.78E-03 |
| ENSG00000105559 | PLEKHA4 | 0.47782949 | 6.05E-03 |
| ENSG00000143387 | CTSK | 0.69479874 | 6.16E-03 |
| ENSG00000118508 | RAB32 | 0.32865418 | 6.17E-03 |
| ENSG00000153071 | DAB2 | 0.45661381 | 6.17E-03 |
| ENSG00000232187 | FTH1P7 | -0.3376602 | 6.24E-03 |
| ENSG00000204856 | FAM216A | 0.25297495 | 6.24E-03 |
| ENSG00000171246 | NPTX1 | -0.623603 | 6.25E-03 |
| ENSG00000169504 | CLIC4 | 0.34258622 | 6.42E-03 |
| ENSG00000129038 | LOXL1 | 0.49987872 | 6.46E-03 |
| ENSG00000072682 | P4HA2 | 0.2964551 | 6.46E-03 |
| ENSG00000104723 | TUSC3 | 0.32786154 | 6.46E-03 |
| ENSG00000010932 | FMO1 | 0.38556968 | 6.46E-03 |
| ENSG00000162722 | TRIM58 | -0.2609433 | 6.46E-03 |
| ENSG00000156218 | ADAMTSL3 | 0.29395277 | 6.46E-03 |
| ENSG00000119699 | TGFB3 | 0.40722164 | 6.46E-03 |
| ENSG00000149212 | SESN3 | 0.44445472 | 6.48E-03 |
| ENSG00000183580 | FBXL7 | 0.31737497 | 6.48E-03 |
| ENSG00000143554 | SLC27A3 | 0.26652326 | 6.48E-03 |
| ENSG00000139354 | GAS2L3 | 0.26526231 | 6.48E-03 |
| ENSG00000142173 | COL6A2 | 0.57265193 | 6.49E-03 |
| ENSG00000136026 | CKAP4 | 0.25564161 | 6.49E-03 |
| ENSG00000121281 | ADCY7 | 0.39521038 | 6.61E-03 |
| ENSG00000000971 | CFH | 0.4259001 | 6.63E-03 |
| ENSG00000101162 | TUBB1 | -0.5617764 | 6.81E-03 |
| ENSG00000072401 | UBE2D1 | -0.2560523 | 6.81E-03 |
| ENSG00000139514 | SLC7A1 | 0.45178612 | 6.83E-03 |
| ENSG00000133083 | DCLK1 | 0.77239669 | 6.83E-03 |
| ENSG00000159251 | ACTC1 | 1.20548278 | 6.86E-03 |
| ENSG00000198795 | ZNF521 | 0.33684315 | 7.15E-03 |
| ENSG00000156219 | ART3 | -0.2687006 | 7.26E-03 |
| ENSG00000117114 | ADGRL2 | 0.30571236 | 7.26E-03 |
| ENSG00000108950 | FAM20A | 0.42621384 | 7.47E-03 |
| ENSG00000186326 | RGS9BP | -0.3499338 | 7.70E-03 |
| ENSG00000106772 | PRUNE2 | 0.88776789 | 7.76E-03 |
| ENSG00000237172 | B3GNT9 | 0.36463554 | 7.77E-03 |
| ENSG00000134020 | PEBP4 | -0.3191892 | 7.80E-03 |
| ENSG00000136059 | VILL | 0.27451976 | 7.80E-03 |
| ENSG00000169604 | ANTXR1 | 0.4625148 | 7.80E-03 |
| ENSG00000083857 | FAT1 | 0.5162555 | 7.97E-03 |

| | | | |
|---|---|---|---|
| ENSG00000082684 | SEMA5B | 0.40392167 | 7.98E-03 |
| ENSG00000170962 | PDGFD | 0.54866201 | 7.98E-03 |
| ENSG00000144857 | BOC | 0.38084365 | 8.00E-03 |
| ENSG00000197483 | ZNF628 | -0.2661205 | 8.02E-03 |
| ENSG00000143061 | IGSF3 | 0.49714363 | 8.02E-03 |
| ENSG00000117155 | SSX2IP | 0.29219857 | 8.07E-03 |
| ENSG00000154721 | JAM2 | 0.32710159 | 8.13E-03 |
| ENSG00000136010 | ALDH1L2 | 0.4251683 | 8.19E-03 |
| ENSG00000183160 | TMEM119 | 0.7101973 | 8.32E-03 |
| ENSG00000184867 | ARMCX2 | 0.26814828 | 8.36E-03 |
| ENSG00000106665 | CLIP2 | 0.26911347 | 8.44E-03 |
| ENSG00000255154 | RP11-80H18.3 | -0.2692621 | 8.45E-03 |
| ENSG00000167210 | LOXHD1 | 0.27739009 | 8.46E-03 |
| ENSG00000088827 | SIGLEC1 | 0.49331661 | 8.49E-03 |
| ENSG00000136235 | GPNMB | 0.39865969 | 8.49E-03 |
| ENSG00000150961 | SEC24D | 0.41789025 | 8.49E-03 |
| ENSG00000136099 | PCDH8 | -0.3648599 | 8.49E-03 |
| ENSG00000172137 | CALB2 | -0.2877653 | 8.53E-03 |
| ENSG00000122644 | ARL4A | 0.3408076 | 8.66E-03 |
| ENSG00000102466 | FGF14 | 0.32564548 | 8.70E-03 |
| ENSG00000106080 | FKBP14 | 0.27549853 | 8.70E-03 |
| ENSG00000123572 | NRK | 0.48249982 | 8.70E-03 |
| ENSG00000176194 | CIDEA | -0.3352093 | 8.77E-03 |
| ENSG00000145536 | ADAMTS16 | 0.40947082 | 8.91E-03 |
| ENSG00000075223 | SEMA3C | 0.33587201 | 8.91E-03 |
| ENSG00000107821 | KAZALD1 | 0.49077862 | 9.17E-03 |
| ENSG00000142552 | RCN3 | 0.42071676 | 9.26E-03 |
| ENSG00000107731 | UNC5B | 0.32166297 | 9.31E-03 |
| ENSG00000141756 | FKBP10 | 0.47766242 | 9.32E-03 |
| ENSG00000144824 | PHLDB2 | 0.36832514 | 9.37E-03 |
| ENSG00000227051 | C14orf132 | 0.42259213 | 9.38E-03 |
| ENSG00000150593 | PDCD4 | 0.25595158 | 9.48E-03 |
| ENSG00000116793 | PHTF1 | 0.26644368 | 9.53E-03 |
| ENSG00000173898 | SPTBN2 | 0.26614009 | 9.64E-03 |
| ENSG00000136160 | EDNRB | 0.39255475 | 9.64E-03 |
| ENSG00000122778 | KIAA1549 | 0.3608274 | 9.83E-03 |
| ENSG00000100196 | KDELR3 | 0.48829017 | 9.83E-03 |
| ENSG00000106823 | ECM2 | 0.51822002 | 9.83E-03 |
| ENSG00000144730 | IL17RD | 0.35402851 | 9.83E-03 |
| ENSG00000117632 | STMN1 | 0.27912237 | 9.83E-03 |
| ENSG00000167552 | TUBA1A | 0.46313392 | 9.83E-03 |
| ENSG00000154175 | ABI3BP | 0.53922306 | 9.83E-03 |

| | | | |
|---|---|---|---|
| ENSG00000106537 | TSPAN13 | 0.2751042 | 9.86E-03 |
| ENSG00000187288 | CIDEC | -0.639777 | 9.90E-03 |
| ENSG00000244405 | ETV5 | 0.43827542 | 9.90E-03 |
| ENSG00000121297 | TSHZ3 | 0.29915409 | 9.91E-03 |

***Supplementary Table 5: Significantly differentially expressed genes in Female Postprandial SAT Samples***

| Ensembl ID | Gene Symbol | log2FC | FDR adjusted P-value |
|---|---|---|---|
| ENSG00000140873 | ADAMTS18 | -0.8935232 | 4.48E-06 |
| ENSG00000147642 | SYBU | 0.25906264 | 1.39E-05 |
| ENSG00000116106 | EPHA4 | 0.46809585 | 1.39E-05 |
| ENSG00000198435 | NRARP | -0.3855681 | 1.81E-05 |
| ENSG00000258316 | KLF17P1 | 0.37183774 | 2.40E-05 |
| ENSG00000153933 | DGKE | 0.30734695 | 2.51E-05 |
| ENSG00000226216 | RPS12P5 | 0.34235903 | 4.06E-05 |
| ENSG00000103044 | HAS3 | -0.4276814 | 4.06E-05 |
| ENSG00000164687 | FABP5 | 0.27001163 | 5.21E-05 |
| ENSG00000152804 | HHEX | -0.3559129 | 5.28E-05 |
| ENSG00000040633 | PHF23 | -0.2669412 | 6.68E-05 |
| ENSG00000164619 | BMPER | 0.39592878 | 7.35E-05 |
| ENSG00000163762 | TM4SF18 | 0.30503074 | 1.10E-04 |
| ENSG00000158406 | HIST1H4H | -0.4773356 | 1.16E-04 |
| ENSG00000168916 | ZNF608 | 0.26915083 | 1.17E-04 |
| ENSG00000260124 | NA | -0.3611081 | 1.20E-04 |
| ENSG00000109107 | ALDOC | -0.472639 | 1.28E-04 |
| ENSG00000172367 | PDZD3 | -0.5175686 | 1.28E-04 |
| ENSG00000111110 | PPM1H | 0.45811325 | 1.28E-04 |
| ENSG00000137713 | PPP2R1B | -0.3525979 | 1.28E-04 |
| ENSG00000113532 | ST8SIA4 | 0.40316002 | 1.36E-04 |
| ENSG00000152689 | RASGRP3 | 0.35621442 | 1.36E-04 |
| ENSG00000132688 | NES | 0.3042809 | 2.10E-04 |
| ENSG00000163874 | ZC3H12A | -0.2881312 | 2.56E-04 |
| ENSG00000149798 | CDC42EP2 | -0.2796822 | 2.56E-04 |
| ENSG00000188522 | FAM83G | -0.2825603 | 2.56E-04 |
| ENSG00000183010 | PYCR1 | 0.41894859 | 2.86E-04 |
| ENSG00000186466 | AQP7P1 | 0.34146819 | 2.95E-04 |
| ENSG00000182580 | EPHB3 | -0.4352545 | 2.95E-04 |
| ENSG00000065054 | SLC9A3R2 | 0.2908976 | 2.95E-04 |
| ENSG00000129422 | MTUS1 | 0.25073141 | 2.95E-04 |
| ENSG00000141854 | MISP3 | -0.3033611 | 2.99E-04 |
| ENSG00000121904 | CSMD2 | 0.45554889 | 2.99E-04 |
| ENSG00000138411 | HECW2 | 0.49091943 | 2.99E-04 |
| ENSG00000019102 | VSIG2 | -0.3560675 | 3.06E-04 |

| | | | |
|---|---|---|---|
| ENSG00000160888 | IER2 | -0.28984 | 3.06E-04 |
| ENSG00000087237 | CETP | 0.80432077 | 3.11E-04 |
| ENSG00000267107 | PCAT19 | 0.26203346 | 3.19E-04 |
| ENSG00000180318 | ALX1 | -0.4308796 | 3.31E-04 |
| ENSG00000163449 | TMEM169 | -0.365346 | 3.31E-04 |
| ENSG00000258545 | RHOXF1-AS1 | -0.3489976 | 3.49E-04 |
| ENSG00000150594 | ADRA2A | -0.2980405 | 3.65E-04 |
| ENSG00000162520 | SYNC | -0.3690873 | 3.73E-04 |
| ENSG00000135636 | DYSF | 0.33093777 | 3.85E-04 |
| ENSG00000187479 | C11orf96 | -0.350815 | 4.13E-04 |
| ENSG00000101955 | SRPX | 0.25370715 | 4.13E-04 |
| ENSG00000131759 | RARA | -0.2615513 | 4.29E-04 |
| ENSG00000145632 | PLK2 | -0.3020078 | 4.61E-04 |
| ENSG00000236882 | LINC01554 | 0.32991895 | 4.64E-04 |
| ENSG00000134363 | FST | 0.31423789 | 4.64E-04 |
| ENSG00000131634 | TMEM204 | 0.25463102 | 4.76E-04 |
| ENSG00000141232 | TOB1 | -0.2544728 | 4.93E-04 |
| ENSG00000138131 | LOXL4 | -0.293501 | 5.98E-04 |
| ENSG00000178826 | TMEM139 | -0.499695 | 6.22E-04 |
| ENSG00000088882 | CPXM1 | 0.47015106 | 6.22E-04 |
| ENSG00000020577 | SAMD4A | -0.3014904 | 6.53E-04 |
| ENSG00000167470 | MIDN | -0.323002 | 6.53E-04 |
| ENSG00000168994 | PXDC1 | -0.2575875 | 6.67E-04 |
| ENSG00000172985 | SH3RF3 | -0.2634729 | 7.16E-04 |
| ENSG00000119138 | KLF9 | 0.25785183 | 7.22E-04 |
| ENSG00000214756 | METTL12 | 0.28479814 | 7.43E-04 |
| ENSG00000169991 | IFFO2 | -0.2758853 | 7.66E-04 |
| ENSG00000158445 | KCNB1 | -0.312515 | 8.49E-04 |
| ENSG00000251390 | NA | -0.3256537 | 9.17E-04 |
| ENSG00000136867 | SLC31A2 | -0.314081 | 9.38E-04 |
| ENSG00000069399 | BCL3 | -0.3431464 | 1.02E-03 |
| ENSG00000187837 | HIST1H1C | -0.3253076 | 1.05E-03 |
| ENSG00000176749 | CDK5R1 | -0.2827619 | 1.09E-03 |
| ENSG00000064989 | CALCRL | 0.34132503 | 1.20E-03 |
| ENSG00000167565 | SERTAD3 | -0.2942758 | 1.21E-03 |
| ENSG00000159640 | ACE | 0.25648476 | 1.31E-03 |
| ENSG00000084710 | EFR3B | 0.36351954 | 1.31E-03 |
| ENSG00000143367 | TUFT1 | -0.3312182 | 1.39E-03 |
| ENSG00000126603 | GLIS2 | -0.3138062 | 1.45E-03 |
| ENSG00000168702 | LRP1B | 0.39949883 | 1.45E-03 |
| ENSG00000180573 | HIST1H2AC | -0.3005266 | 1.50E-03 |
| ENSG00000104267 | CA2 | 0.35547588 | 1.51E-03 |

| | | | |
|---|---|---|---|
| ENSG00000135407 | AVIL | -0.3493123 | 1.55E-03 |
| ENSG00000120068 | HOXB8 | -0.2706089 | 1.62E-03 |
| ENSG00000246430 | LINC00968 | -0.5513061 | 1.63E-03 |
| ENSG00000204389 | HSPA1A | 0.28101828 | 1.69E-03 |
| ENSG00000213613 | RP11-380G5.3 | 0.53613878 | 1.82E-03 |
| ENSG00000183307 | CECR6 | -0.2931439 | 1.87E-03 |
| ENSG00000143171 | RXRG | 0.34712508 | 1.92E-03 |
| ENSG00000128567 | PODXL | 0.25575756 | 1.92E-03 |
| ENSG00000116285 | ERRFI1 | 0.33952701 | 1.94E-03 |
| ENSG00000269736 | CTD-2521M24.11 | -0.3564901 | 1.96E-03 |
| ENSG00000184545 | DUSP8 | -0.3574684 | 2.11E-03 |
| ENSG00000176533 | GNG7 | 0.27202879 | 2.12E-03 |
| ENSG00000185100 | ADSSL1 | 0.3279583 | 2.22E-03 |
| ENSG00000145283 | SLC10A6 | 0.38234743 | 2.28E-03 |
| ENSG00000179094 | PER1 | 0.33173425 | 2.28E-03 |
| ENSG00000129474 | AJUBA | -0.2893356 | 2.28E-03 |
| ENSG00000234167 | TCEB2P4 | 0.28429966 | 2.45E-03 |
| ENSG00000265635 | MIR3612 | 0.30125727 | 2.51E-03 |
| ENSG00000172216 | CEBPB | -0.3388099 | 2.51E-03 |
| ENSG00000116833 | NR5A2 | 0.27371301 | 2.53E-03 |
| ENSG00000262194 | NA | -0.2809058 | 2.66E-03 |
| ENSG00000107831 | FGF8 | -0.4492058 | 2.76E-03 |
| ENSG00000174236 | REP15 | 0.31507406 | 2.81E-03 |
| ENSG00000257556 | RP11-44N21.1 | -0.3024917 | 2.82E-03 |
| ENSG00000260837 | RP11-434B12.1 | -0.2607968 | 2.87E-03 |
| ENSG00000101665 | SMAD7 | 0.3060424 | 2.87E-03 |
| ENSG00000221340 | RNU6ATAC18P | 0.33666925 | 2.95E-03 |
| ENSG00000189350 | FAM179A | -0.3315763 | 3.07E-03 |
| ENSG00000135441 | BLOC1S1 | -0.3080779 | 3.12E-03 |
| ENSG00000140961 | OSGIN1 | -0.3248109 | 3.12E-03 |
| ENSG00000163638 | ADAMTS9 | 0.29807235 | 3.13E-03 |
| ENSG00000113070 | HBEGF | 0.28152136 | 3.13E-03 |
| ENSG00000164093 | PITX2 | -0.3124456 | 3.21E-03 |
| ENSG00000176236 | C10orf111 | -0.3316134 | 3.25E-03 |
| ENSG00000224939 | LINC00184 | -0.4194841 | 3.26E-03 |
| ENSG00000101883 | RHOXF1 | -0.298862 | 3.30E-03 |
| ENSG00000136492 | BRIP1 | 0.28106493 | 3.39E-03 |
| ENSG00000173898 | SPTBN2 | 0.40643384 | 3.48E-03 |
| ENSG00000234964 | FABP5P7 | 0.25106107 | 3.48E-03 |
| ENSG00000165659 | NA | 0.31979077 | 3.84E-03 |
| ENSG00000198848 | CES1 | -0.3804954 | 3.95E-03 |
| ENSG00000154127 | UBASH3B | 0.3446007 | 3.95E-03 |

| | | | |
|---|---|---|---|
| ENSG00000162426 | SLC45A1 | 0.29169968 | 3.95E-03 |
| ENSG00000165084 | C8orf34 | 0.33331981 | 4.13E-03 |
| ENSG00000165507 | C10orf10 | 0.52981565 | 4.24E-03 |
| ENSG00000182487 | NCF1B | -0.4498776 | 4.29E-03 |
| ENSG00000239622 | CTA-242H14.1 | -0.3389626 | 4.32E-03 |
| ENSG00000117318 | ID3 | -0.2910112 | 4.32E-03 |
| ENSG00000139880 | CDH24 | 0.28063347 | 4.33E-03 |
| ENSG00000138678 | GPAT3 | 0.62214846 | 4.54E-03 |
| ENSG00000260792 | RP11-982M15.8 | -0.308797 | 4.54E-03 |
| ENSG00000228695 | CES1P1 | -0.3565975 | 4.54E-03 |
| ENSG00000029153 | ARNTL2 | 0.29468334 | 4.56E-03 |
| ENSG00000164683 | HEY1 | 0.26823323 | 4.56E-03 |
| ENSG00000132139 | NA | -0.4567237 | 4.57E-03 |
| ENSG00000125414 | MYH2 | -0.4350872 | 4.73E-03 |
| ENSG00000196739 | COL27A1 | 0.27848858 | 4.77E-03 |
| ENSG00000081277 | PKP1 | -0.3534734 | 4.85E-03 |
| ENSG00000099957 | P2RX6 | -0.5248829 | 4.88E-03 |
| ENSG00000180596 | HIST1H2BC | -0.3593388 | 4.89E-03 |
| ENSG00000116133 | DHCR24 | -0.2679734 | 5.11E-03 |
| ENSG00000228594 | FNDC10 | -0.340536 | 5.29E-03 |
| ENSG00000188766 | SPRED3 | -0.2706287 | 5.32E-03 |
| ENSG00000119630 | PGF | -0.3114995 | 5.63E-03 |
| ENSG00000092200 | RPGRIP1 | -0.284768 | 5.87E-03 |
| ENSG00000172572 | PDE3A | 0.28785251 | 5.99E-03 |
| ENSG00000224411 | HSP90AA2P | 0.27944227 | 5.99E-03 |
| ENSG00000221643 | SNORA77 | -0.2741975 | 6.42E-03 |
| ENSG00000197165 | SULT1A2 | -0.4215175 | 6.62E-03 |
| ENSG00000130203 | APOE | 0.40932591 | 6.90E-03 |
| ENSG00000267278 | MAP3K14-AS1 | -0.2816703 | 7.15E-03 |
| ENSG00000188483 | IER5L | -0.2957055 | 7.17E-03 |
| ENSG00000095739 | BAMBI | -0.3063023 | 7.26E-03 |
| ENSG00000244734 | HBB | -0.9572104 | 7.53E-03 |
| ENSG00000152580 | IGSF10 | 0.29091731 | 7.82E-03 |
| ENSG00000236493 | EIF2S2P3 | -0.2674602 | 7.92E-03 |
| ENSG00000105852 | PON3 | 0.25526295 | 8.14E-03 |
| ENSG00000115556 | PLCD4 | 0.29999581 | 8.38E-03 |
| ENSG00000206172 | HBA1 | -0.8533706 | 8.38E-03 |
| ENSG00000136367 | ZFHX2 | 0.31920966 | 8.38E-03 |
| ENSG00000116741 | RGS2 | -0.4162533 | 8.38E-03 |
| ENSG00000163884 | KLF15 | 0.27487375 | 8.38E-03 |
| ENSG00000158104 | HPD | -0.3830906 | 8.47E-03 |
| ENSG00000147883 | CDKN2B | -0.2608329 | 8.98E-03 |

| | | | |
|---|---|---|---|
| ENSG00000170667 | RASA4B | -0.3012842 | 8.99E-03 |
| ENSG00000255449 | RP11-91P24.6 | 0.27974372 | 9.04E-03 |
| ENSG00000153993 | SEMA3D | 0.31804678 | 9.18E-03 |
| ENSG00000262601 | NA | 0.27245135 | 9.37E-03 |
| ENSG00000118971 | CCND2 | -0.2905954 | 9.44E-03 |
| ENSG00000122966 | CIT | -0.3590407 | 9.50E-03 |
| ENSG00000225026 | AC091492.2 | 0.29276259 | 9.59E-03 |

***Supplementary Table 6: Significantly differentially expressed genes in Female Postprandial Muscle Samples***

| Ensembl ID | Gene Symbol | log2FC | FDR adjusted P-value |
|---|---|---|---|
| ENSG00000019549 | SNAI2 | 0.62237737 | 1.33E-04 |
| ENSG00000118263 | KLF7 | 0.45581952 | 7.19E-04 |
| ENSG00000176641 | RNF152 | 0.57281565 | 1.02E-03 |
| ENSG00000079102 | RUNX1T1 | 0.5058928 | 1.09E-03 |
| ENSG00000214717 | ZBED1 | -0.2997235 | 1.09E-03 |
| ENSG00000100478 | AP4S1 | -0.2742075 | 2.54E-03 |
| ENSG00000197283 | SYNGAP1 | 0.56084316 | 2.54E-03 |
| ENSG00000169894 | MUC3A | 0.55211087 | 2.54E-03 |
| ENSG00000169733 | RFNG | -0.2508898 | 2.84E-03 |
| ENSG00000134954 | ETS1 | 0.33130708 | 2.84E-03 |
| ENSG00000075240 | GRAMD4 | 0.32913128 | 2.87E-03 |
| ENSG00000150760 | DOCK1 | 0.41433397 | 3.58E-03 |
| ENSG00000128590 | DNAJB9 | -0.2528533 | 3.58E-03 |
| ENSG00000177666 | PNPLA2 | -0.2697835 | 3.58E-03 |
| ENSG00000215788 | TNFRSF25 | 0.49974531 | 4.14E-03 |
| ENSG00000064309 | CDON | 0.47928318 | 4.14E-03 |
| ENSG00000168916 | ZNF608 | 0.43703353 | 4.14E-03 |
| ENSG00000219481 | NBPF1 | 0.34291646 | 4.14E-03 |
| ENSG00000110274 | CEP164 | 0.40124933 | 4.14E-03 |
| ENSG00000268518 | CTD-2545M3.8 | 0.75326751 | 4.14E-03 |
| ENSG00000130158 | DOCK6 | 0.30126226 | 4.14E-03 |
| ENSG00000204248 | COL11A2 | 0.47758407 | 4.14E-03 |
| ENSG00000088882 | CPXM1 | 1.14267414 | 4.27E-03 |
| ENSG00000096080 | MRPS18A | -0.2545498 | 4.58E-03 |
| ENSG00000106819 | ASPN | 0.77937112 | 4.91E-03 |
| ENSG00000130312 | MRPL34 | -0.3175991 | 4.91E-03 |
| ENSG00000109689 | STIM2 | 0.26662274 | 4.91E-03 |
| ENSG00000183722 | LHFP | 0.55688195 | 4.91E-03 |
| ENSG00000132122 | SPATA6 | 0.38125475 | 4.91E-03 |
| ENSG00000106511 | MEOX2 | 0.39477034 | 4.91E-03 |
| ENSG00000084636 | COL16A1 | 0.5515381 | 4.91E-03 |
| ENSG00000166192 | SENP8 | -0.2999922 | 4.91E-03 |
| ENSG00000145014 | TMEM44 | 0.34148207 | 4.91E-03 |
| ENSG00000174080 | CTSF | -0.305075 | 4.91E-03 |
| ENSG00000128641 | MYO1B | 0.36678368 | 4.91E-03 |
| ENSG00000156504 | FAM122B | 0.31123816 | 4.91E-03 |
| ENSG00000181817 | LSM10 | -0.259757 | 4.91E-03 |
| ENSG00000204262 | COL5A2 | 0.8930227 | 4.91E-03 |

| | | | |
|---|---|---|---|
| ENSG00000154783 | FGD5 | 0.32742345 | 4.91E-03 |
| ENSG00000111145 | ELK3 | 0.34634159 | 4.91E-03 |
| ENSG00000165898 | ISCA2 | -0.2572494 | 4.91E-03 |
| ENSG00000267480 | RP11-703I16.1 | -0.3605915 | 4.91E-03 |
| ENSG00000151779 | NBAS | -0.3106099 | 4.91E-03 |
| ENSG00000214021 | TTLL3 | 0.46022941 | 4.91E-03 |
| ENSG00000103335 | PIEZO1 | 0.43258298 | 4.91E-03 |
| ENSG00000163565 | IFI16 | 0.42000772 | 4.91E-03 |
| ENSG00000106823 | ECM2 | 0.4784313 | 4.91E-03 |
| ENSG00000081923 | ATP8B1 | 0.33508548 | 4.94E-03 |
| ENSG00000260336 | NA | 0.32894472 | 4.95E-03 |
| ENSG00000118495 | PLAGL1 | 0.46575285 | 5.03E-03 |
| ENSG00000090924 | PLEKHG2 | 0.48704336 | 5.07E-03 |
| ENSG00000112139 | MDGA1 | 0.46970164 | 5.19E-03 |
| ENSG00000139625 | MAP3K12 | 0.4266771 | 5.25E-03 |
| ENSG00000198795 | ZNF521 | 0.3961381 | 5.25E-03 |
| ENSG00000105287 | PRKD2 | 0.28774804 | 5.25E-03 |
| ENSG00000111961 | SASH1 | 0.25631561 | 5.25E-03 |
| ENSG00000248923 | MTND5P11 | -0.4312959 | 5.34E-03 |
| ENSG00000221818 | EBF2 | 0.29678551 | 5.34E-03 |
| ENSG00000135540 | NHSL1 | 0.41179493 | 5.42E-03 |
| ENSG00000135766 | EGLN1 | -0.3166438 | 5.44E-03 |
| ENSG00000171992 | SYNPO | -0.3494241 | 5.44E-03 |
| ENSG00000117594 | HSD11B1 | -0.5059585 | 5.47E-03 |
| ENSG00000147872 | PLIN2 | -0.3169415 | 5.48E-03 |
| ENSG00000164741 | DLC1 | 0.26979428 | 5.48E-03 |
| ENSG00000241399 | CD302 | 0.30000782 | 5.71E-03 |
| ENSG00000082196 | C1QTNF3 | 0.44094218 | 5.75E-03 |
| ENSG00000139679 | LPAR6 | 0.31897593 | 5.80E-03 |
| ENSG00000116663 | FBXO6 | -0.3308512 | 5.80E-03 |
| ENSG00000074696 | HACD3 | 0.33540476 | 5.86E-03 |
| ENSG00000237461 | RP11-554F20.1 | -0.2670253 | 5.89E-03 |
| ENSG00000214942 | AC113167.1 | -0.3505529 | 5.94E-03 |
| ENSG00000171680 | PLEKHG5 | 0.45549928 | 5.94E-03 |
| ENSG00000106546 | AHR | 0.34484942 | 5.96E-03 |
| ENSG00000126785 | RHOJ | 0.30144469 | 5.96E-03 |
| ENSG00000101400 | SNTA1 | -0.3079476 | 5.96E-03 |
| ENSG00000010810 | FYN | 0.28464756 | 5.96E-03 |
| ENSG00000143387 | CTSK | 0.54232653 | 5.96E-03 |
| ENSG00000125844 | RRBP1 | 0.36062288 | 5.96E-03 |
| ENSG00000069535 | MAOB | -0.2943647 | 6.13E-03 |
| ENSG00000168487 | BMP1 | 0.51728459 | 6.16E-03 |

| | | | |
|---|---|---|---|
| ENSG00000137338 | PGBD1 | 0.27374612 | 6.20E-03 |
| ENSG00000066056 | TIE1 | 0.3156074 | 6.38E-03 |
| ENSG00000080573 | COL5A3 | 0.64198399 | 6.38E-03 |
| ENSG00000130702 | LAMA5 | 0.39287834 | 6.38E-03 |
| ENSG00000108387 | Sep/04 | 0.34406262 | 6.38E-03 |
| ENSG00000264868 | NA | 0.42516668 | 6.38E-03 |
| ENSG00000136378 | ADAMTS7 | 0.57196492 | 6.81E-03 |
| ENSG00000085276 | MECOM | 0.35192443 | 6.81E-03 |
| ENSG00000108821 | COL1A1 | 1.38250321 | 7.22E-03 |
| ENSG00000160360 | GPSM1 | 0.39600316 | 7.22E-03 |
| ENSG00000118257 | NRP2 | 0.39902425 | 7.22E-03 |
| ENSG00000143515 | ATP8B2 | 0.35663923 | 7.22E-03 |
| ENSG00000061273 | HDAC7 | 0.31481134 | 7.42E-03 |
| ENSG00000162367 | TAL1 | 0.3856787 | 7.45E-03 |
| ENSG00000154721 | JAM2 | 0.30718678 | 7.45E-03 |
| ENSG00000169683 | LRRC45 | 0.28264506 | 7.45E-03 |
| ENSG00000184500 | PROS1 | 0.29054506 | 7.45E-03 |
| ENSG00000198805 | PNP | 0.39323477 | 7.45E-03 |
| ENSG00000111245 | MYL2 | -0.3795041 | 7.59E-03 |
| ENSG00000179632 | MAF1 | -0.2792151 | 7.83E-03 |
| ENSG00000147642 | SYBU | 0.29785073 | 7.84E-03 |
| ENSG00000164099 | PRSS12 | 0.54446431 | 7.84E-03 |
| ENSG00000069020 | MAST4 | 0.29676688 | 7.84E-03 |
| ENSG00000100968 | NFATC4 | 0.37303496 | 7.84E-03 |
| ENSG00000159433 | STARD9 | 0.42039813 | 7.92E-03 |
| ENSG00000139117 | CPNE8 | 0.33785589 | 7.92E-03 |
| ENSG00000171813 | PWWP2B | -0.3094081 | 8.07E-03 |
| ENSG00000107731 | UNC5B | 0.35475356 | 8.35E-03 |
| ENSG00000225526 | MKRN2OS | -0.2961963 | 8.52E-03 |
| ENSG00000167766 | ZNF83 | 0.44948567 | 8.52E-03 |
| ENSG00000145198 | VWA5B2 | -0.304449 | 8.52E-03 |
| ENSG00000204301 | NOTCH4 | 0.35997222 | 8.52E-03 |
| ENSG00000182796 | TMEM198B | 0.41876159 | 8.61E-03 |
| ENSG00000117519 | CNN3 | 0.26785378 | 8.71E-03 |
| ENSG00000118971 | CCND2 | 0.42840136 | 8.71E-03 |
| ENSG00000204427 | ABHD16A | -0.3232531 | 8.71E-03 |
| ENSG00000204681 | GABBR1 | 0.59506439 | 8.85E-03 |
| ENSG00000181045 | SLC26A11 | 0.32349621 | 8.88E-03 |
| ENSG00000072071 | ADGRL1 | 0.32737174 | 8.91E-03 |
| ENSG00000087903 | RFX2 | 0.40727424 | 8.91E-03 |
| ENSG00000091592 | NLRP1 | 0.36196907 | 8.91E-03 |
| ENSG00000161381 | PLXDC1 | 0.3974883 | 8.91E-03 |

| | | | |
|---|---|---|---|
| ENSG00000130635 | COL5A1 | 0.84256519 | 9.08E-03 |
| ENSG00000188130 | MAPK12 | -0.2562858 | 9.08E-03 |
| ENSG00000004478 | FKBP4 | -0.274058 | 9.17E-03 |
| ENSG00000166130 | IKBIP | 0.34316445 | 9.17E-03 |
| ENSG00000108797 | CNTNAP1 | 0.38655585 | 9.23E-03 |
| ENSG00000159069 | FBXW5 | -0.2615027 | 9.42E-03 |
| ENSG00000137857 | DUOX1 | 0.33250851 | 9.43E-03 |
| ENSG00000101695 | RNF125 | 0.3773887 | 9.43E-03 |
| ENSG00000125037 | EMC3 | -0.2657663 | 9.43E-03 |
| ENSG00000153317 | ASAP1 | 0.3517889 | 9.43E-03 |
| ENSG00000138594 | TMOD3 | 0.25245345 | 9.43E-03 |
| ENSG00000108861 | DUSP3 | -0.272503 | 9.43E-03 |
| ENSG00000196562 | SULF2 | 0.34077964 | 9.43E-03 |
| ENSG00000052126 | PLEKHA5 | 0.3577237 | 9.43E-03 |
| ENSG00000159348 | CYB5R1 | -0.262379 | 9.43E-03 |
| ENSG00000112530 | PACRG | -0.3284224 | 9.43E-03 |
| ENSG00000160305 | DIP2A | 0.33447931 | 9.43E-03 |
| ENSG00000018408 | WWTR1 | 0.27699556 | 9.43E-03 |
| ENSG00000091136 | LAMB1 | 0.41015429 | 9.43E-03 |
| ENSG00000168542 | COL3A1 | 1.06343132 | 9.43E-03 |
| ENSG00000123560 | PLP1 | 0.59408274 | 9.43E-03 |
| ENSG00000134802 | SLC43A3 | 0.32219937 | 9.43E-03 |
| ENSG00000160796 | NBEAL2 | 0.38141076 | 9.50E-03 |
| ENSG00000260806 | RP11-872J21.3 | -0.2757424 | 9.50E-03 |
| ENSG00000179262 | RAD23A | -0.3175492 | 9.50E-03 |
| ENSG00000185813 | PCYT2 | -0.3343442 | 9.50E-03 |
| ENSG00000142409 | ZNF787 | -0.2743578 | 9.50E-03 |
| ENSG00000112245 | PTP4A1 | -0.2787245 | 9.50E-03 |
| ENSG00000169231 | THBS3 | 0.44447831 | 9.50E-03 |
| ENSG00000106868 | SUSD1 | 0.31259686 | 9.50E-03 |
| ENSG00000124593 | PRICKLE4 | 0.38422906 | 9.50E-03 |
| ENSG00000175274 | TP53I11 | 0.35601394 | 9.50E-03 |
| ENSG00000239857 | GET4 | 0.35231847 | 9.50E-03 |
| ENSG00000164330 | EBF1 | 0.30633594 | 9.50E-03 |
| ENSG00000049130 | KITLG | 0.31078883 | 9.50E-03 |
| ENSG00000257859 | CASC18 | -0.3755866 | 9.50E-03 |
| ENSG00000175084 | DES | -0.2767186 | 9.56E-03 |
| ENSG00000103994 | ZNF106 | -0.2551654 | 9.56E-03 |
| ENSG00000151067 | CACNA1C | 0.33068955 | 9.59E-03 |
| ENSG00000077274 | CAPN6 | 0.77404905 | 9.59E-03 |
| ENSG00000129467 | ADCY4 | 0.41472781 | 9.59E-03 |
| ENSG00000133247 | KMT5C | 0.36038293 | 9.66E-03 |

| | | | |
|---|---|---|---|
| ENSG00000172137 | CALB2 | -0.3582839 | 9.76E-03 |
| ENSG00000099364 | FBXL19 | 0.28101707 | 9.78E-03 |
| ENSG00000127423 | AUNIP | -0.3276192 | 9.87E-03 |
| ENSG00000184584 | TMEM173 | 0.31646583 | 9.91E-03 |

***Supplementary Table 7: Overview of the genes involved in the pathway enrichment***

| Tissue | Sex | Pathway | Adjusted P-value | N pathway genes expressed | Significantly downregulated DEGs | Significantly upregulated DEGs |
|---|---|---|---|---|---|---|
| Postprandial SAT Transcriptome | Male Samples | Fertilization | 2.54E-03 | 15 | ACR CATSPERG | ZP1 |
| Postprandial SAT Transcriptome | Male Samples | Binding and Uptake of Ligands by Scavenger Receptors | 3.27E-03 | 46 | SPARC HBB | APOE HSPH1 |
| Postprandial SAT Transcriptome | Male Samples | Signaling by Nuclear Receptors | 2.12E-02 | 214 | CYP26B1 | APOE APOC1 RXRG CETP FKBP4 |
| Postprandial SAT Transcriptome | Male Samples | HDL remodeling | 2.94E-02 | 7 | | APOE CETP |
| Postprandial SAT Transcriptome | Male Samples | NR1H3 & NR1H2 regulate gene expression linked to cholesterol transport and efflux | 3.74E-02 | 36 | | APOE APOC1 CETP |
| Postprandial Muscle Transcriptome | Male Samples | Extracellular matrix organization | 5.32E-24 | 241 | | NID2 COL1A1 COL3A1 SERPINH1 ADAMTS2 CAPN6 COL5A2 COL1A2 PXDN COL4A1 COL5A1 LOXL2 P3H1 COL14A1 COL4A2 ASPN LAMA4 COL6A6 COL6A3 LOX ELN SPARC COL15A1 NID1 LAMB1 COL6A1 TGFB2 LUM BGN HTRA1 MMP2 CTSK LOXL1 P4HA2 TGFB3 COL6A2 JAM2 ADAMTS16 |
| Postprandial Muscle Transcriptome | Male Samples | Collagen formation | 8.31E-18 | 80 | | COL1A1 COL3A1 SERPINH1 ADAMTS2 COL5A2 COL1A2 PXDN COL4A1 COL5A1 LOXL2 P3H1 COL14A1 COL4A2 COL6A6 COL6A3 LOX COL15A1 COL6A1 LOXL1 P4HA2 COL6A2 |
| Postprandial Muscle Transcriptome | Male Samples | Assembly of collagen fibrils and other multimeric structures | 4.1E-15 | 49 | | COL1A1 COL3A1 COL5A2 COL1A2 PXDN COL4A1 COL5A1 LOXL2 COL4A2 COL6A6 COL6A3 LOX COL15A1 COL6A1 LOXL1 COL6A2 |
| Postprandial Muscle Transcriptome | Male Samples | Collagen biosynthesis and modifying enzymes | 7.22E-15 | 61 | | COL1A1 COL3A1 SERPINH1 ADAMTS2 COL5A2 COL1A2 COL4A1 COL5A1 P3H1 COL14A1 COL4A2 COL6A6 COL6A3 COL15A1 COL6A1 P4HA2 COL6A2 |
| Postprandial Muscle Transcriptome | Male Samples | Collagen chain trimerization | 9.88E-13 | 38 | | COL1A1 COL3A1 COL5A2 COL1A2 COL4A1 COL5A1 COL14A1 COL4A2 COL6A6 |

| | | | | | | |
|---|---|---|---|---|---|---|
| | | | | | | COL6A3 COL15A1 COL6A1 COL6A2 |
| Postprandial Muscle Transcriptome | Male Samples | Binding and Uptake of Ligands by Scavenger Receptors | 6.92E-07 | 46 | HBA1 HBB | COL1A1 COL3A1 COL1A2 COL4A1 COL4A2 SPARC COLEC12 |
| Postprandial Muscle Transcriptome | Male Samples | Scavenging by Class A Receptors | 1.24E-05 | 17 | | COL1A1 COL3A1 COL1A2 COL4A1 COL4A2 COLEC12 |
| Postprandial Muscle Transcriptome | Male Samples | ECM proteoglycans | 3.81E-05 | 44 | | ASPN LAMA4 SPARC LAMB1 TGFB2 LUM BGN TGFB3 |
| Postprandial Muscle Transcriptome | Male Samples | Signaling by PDGF | 6.45E-05 | 47 | | THBS4 COL4A1 COL4A2 COL6A6 COL6A3 COL6A1 COL6A2 PDGFD |
| Postprandial Muscle Transcriptome | Male Samples | NCAM1 interactions | 2.55E-04 | 27 | | COL4A1 COL4A2 COL6A6 COL6A3 COL6A1 COL6A2 |
| Postprandial Muscle Transcriptome | Male Samples | Degradation of the extracellular matrix | 6.18E-04 | 82 | | CAPN6 COL14A1 COL15A1 NID1 LAMB1 HTRA1 MMP2 CTSK ADAMTS16 |
| Postprandial Muscle Transcriptome | Male Samples | Crosslinking of collagen fibrils | 6.97E-04 | 10 | | PXDN LOXL2 LOX LOXL1 |
| Postprandial Muscle Transcriptome | Male Samples | NCAM signaling for neurite out-growth | 8.27E-04 | 48 | | COL4A1 COL4A2 COL6A6 COL6A3 COL6A1 COL6A2 SPTBN2 |
| Postprandial Muscle Transcriptome | Male Samples | Elastic fibre formation | 1.46E-03 | 36 | | LOXL2 LOX ELN TGFB2 LOXL1 TGFB3 |
| Postprandial Muscle Transcriptome | Male Samples | Defective B3GALTL causes PpS | 1.18E-02 | 34 | | ADAMTS2 ADAMTS7 ADAMTSL3 SEMA5B ADAMTS16 |
| Postprandial Muscle Transcriptome | Male Samples | O-glycosylation of TSR domain-containing proteins | 1.36E-02 | 35 | | ADAMTS2 ADAMTS7 ADAMTSL3 SEMA5B ADAMTS16 |
| Postprandial Muscle Transcriptome | Male Samples | Laminin interactions | 2.04E-02 | 22 | | NID2 LAMA4 NID1 LAMB1 |
| Postprandial Muscle Transcriptome | Male Samples | Collagen degradation | 3.93E-02 | 26 | | COL14A1 COL15A1 MMP2 CTSK |
| Postprandial SAT Transcriptome | Female Samples | Erythrocytes take up oxygen and release carbon dioxide | 7.86E-05 | 6 | HBB HBA1 | CA2 |
| Postprandial SAT Transcriptome | Female Samples | Erythrocytes take up carbon dioxide and release oxygen | 4.6E-04 | 10 | HBB HBA1 | CA2 |
| Postprandial SAT Transcriptome | Female Samples | O2/CO2 exchange in erythrocytes | 4.6E-04 | 10 | HBB HBA1 | CA2 |

| | | | | | | |
|---|---|---|---|---|---|---|
| Postprandial SAT Transcriptome | Female Samples | Signaling by Nuclear Receptors | 2.76E-03 | 214 | RARA | FABP5 CETP RXRG GNG7 NR5A2 HBEGF APOE |
| Postprandial SAT Transcriptome | Female Samples | HDL remodeling | 9.99E-03 | 7 | | CETP APOE |
| Postprandial SAT Transcriptome | Female Samples | Signal Transduction | 1.56E-02 | 2099 | PPP2R1B CDC42EP2 ADRA2A RARA CDK5R1 DUSP8 FGF8 ID3 SPRED3 PGF BAMBI RGS2 CDKN2B CIT | DGKE FABP5 RASGRP3 CETP FST CALCRL RXRG GNG7 NR5A2 SMAD7 HBEGF SPTBN2 HEY1 PDE3A APOE |
| Postprandial SAT Transcriptome | Female Samples | MAPK family signaling cascades | 1.62E-02 | 280 | PPP2R1B CDC42EP2 DUSP8 FGF8 SPRED3 | RASGRP3 HBEGF SPTBN2 |
| Postprandial Muscle Transcriptome | Female Samples | Collagen chain trimerization | 2.67E-06 | 38 | | COL11A2 COL16A1 COL5A2 COL5A3 COL1A1 COL5A1 COL3A1 |
| Postprandial Muscle Transcriptome | Female Samples | Collagen biosynthesis and modifying enzymes | 5.73E-06 | 61 | | COL11A2 COL16A1 COL5A2 BMP1 COL5A3 COL1A1 COL5A1 COL3A1 |
| Postprandial Muscle Transcriptome | Female Samples | Extracellular matrix organization | 1.11E-05 | 241 | | COL11A2 ASPN COL16A1 COL5A2 CTSK BMP1 COL5A3 LAMA5 COL1A1 JAM2 COL5A1 LAMB1 COL3A1 CAPN6 |
| Postprandial Muscle Transcriptome | Female Samples | Assembly of collagen fibrils and other multimeric structures | 1.62E-05 | 49 | | COL11A2 COL5A2 BMP1 COL5A3 COL1A1 COL5A1 COL3A1 |
| Postprandial Muscle Transcriptome | Female Samples | Collagen formation | 4.65E-05 | 80 | | COL11A2 COL16A1 COL5A2 BMP1 COL5A3 COL1A1 COL5A1 COL3A1 |
| Postprandial Muscle Transcriptome | Female Samples | Degradation of the extracellular matrix | 4.07E-03 | 82 | | COL16A1 CTSK BMP1 LAMA5 LAMB1 CAPN6 |
| Postprandial Muscle Transcriptome | Female Samples | Netrin-1 signaling | 1.38E-02 | 42 | MAPK12 | DOCK1 FYN UNC5B |
| Postprandial Muscle Transcriptome | Female Samples | RUNX2 regulates osteoblast differentiation | 1.91E-02 | 22 | | ZNF521 COL1A1 WWTR1 |

***Supplementary Table 8: Rank overview of JIVE decomposition in male and female samples.***

| | Male Samples | Female Samples |
|---|---|---|
| Joint rank | 2 | 0 |
| Blood individual rank | 11 | 7 |
| SAT individual rank | 15 | 12 |
| Muscle individual rank | 15 | 8 |

### Supplementary Figures

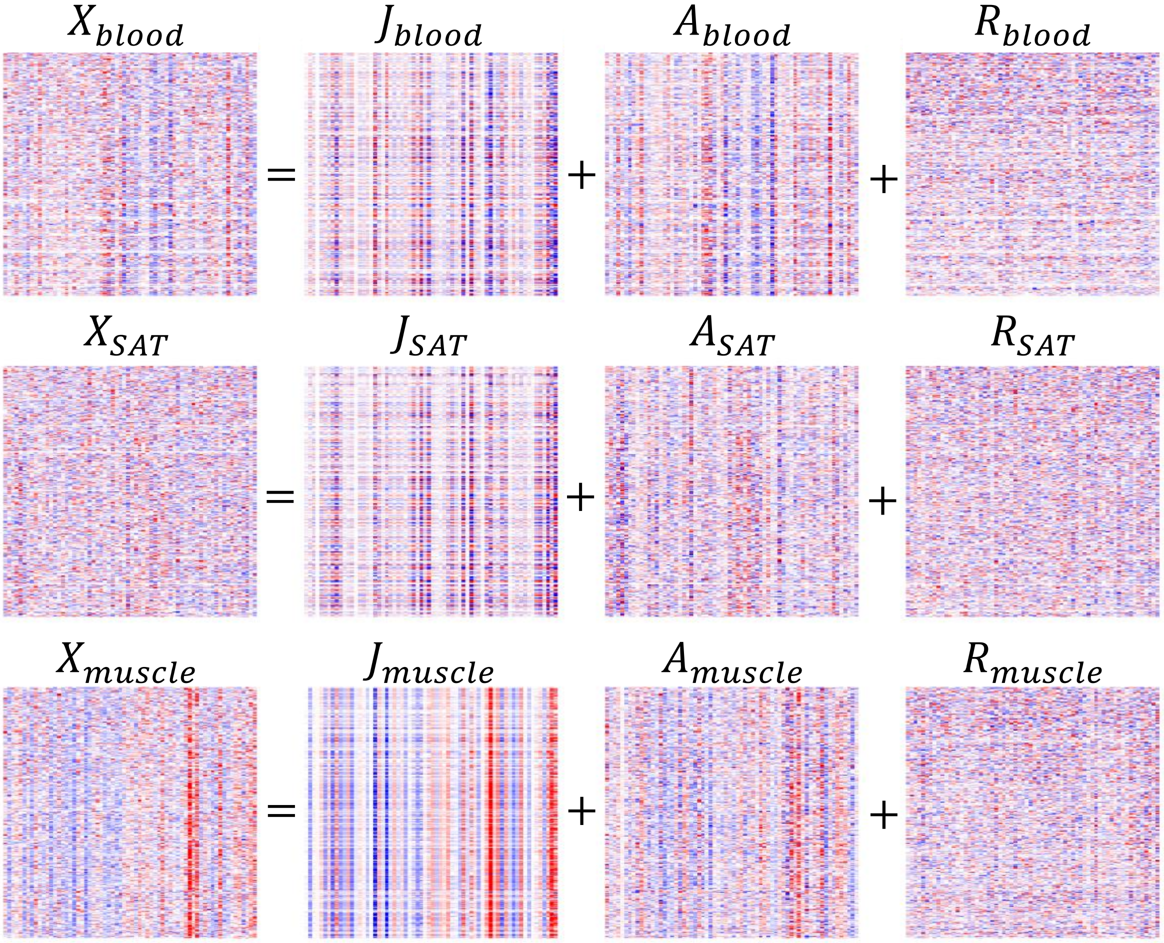


***Supplementary Figure 1: Schematic overview of the JIVE decomposition of male postprandial blood, SAT and muscle transcriptome data.*** *In short, the input expression data of blood, SAT and muscle (X) is decomposed into three matrices: a joint effect (J), individual effect (A), and residual variation (R). Rows represent genes and columns represent samples.* $J_{blood}$, $J_{SAT}$ *and* $J_{muscle}$ *represent a shared variation across the three tissues that is linearly independent from the tissue specific individual effects* $A_{blood}$, $A_{SAT}$ *and* $A_{muscle}$. $R_{blood}$, $R_{SAT}$ *and* $R_{muscle}$ *represent variation that cannot be mapped to either J or A. Blue corresponds to negative values and red to positive values.*

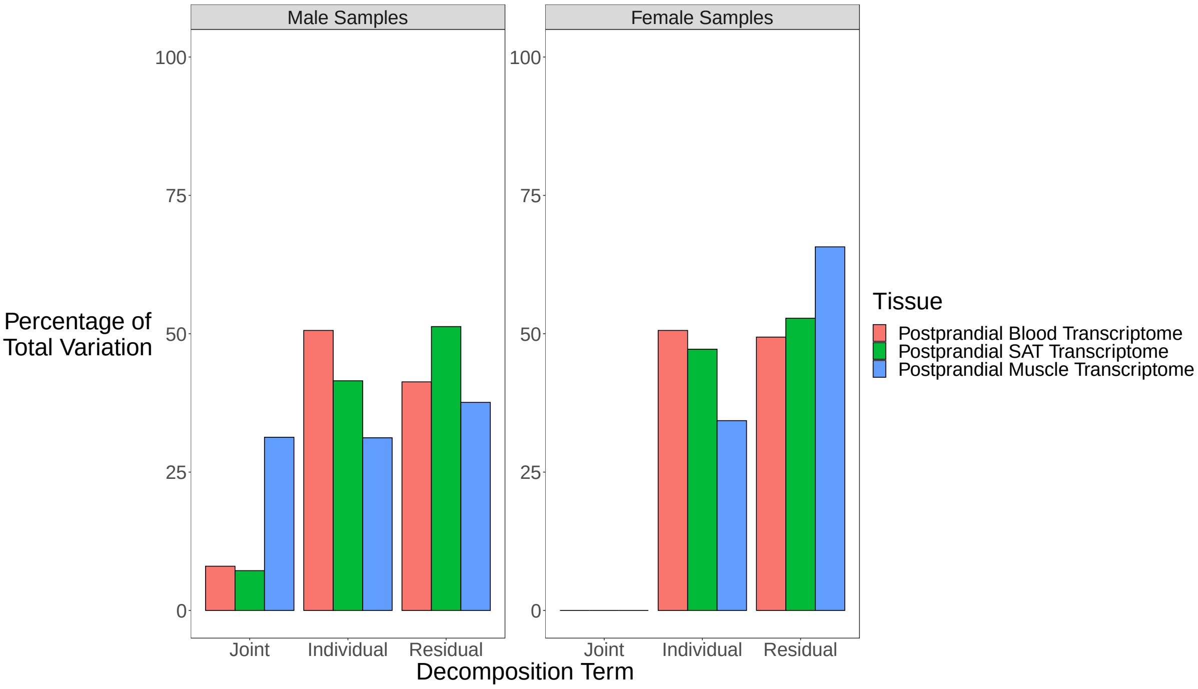


***Supplementary Figure 2: Only male samples have a joint effect across the postprandial blood, SAT and muscle transcriptome****. X-axis represents the decomposition terms. Y-axis represents the percentage of total variation per tissue. Colors represent different tissues; red: postprandial blood transcriptome, green: postprandial SAT transcriptome, blue: postprandial muscle transcriptome.*

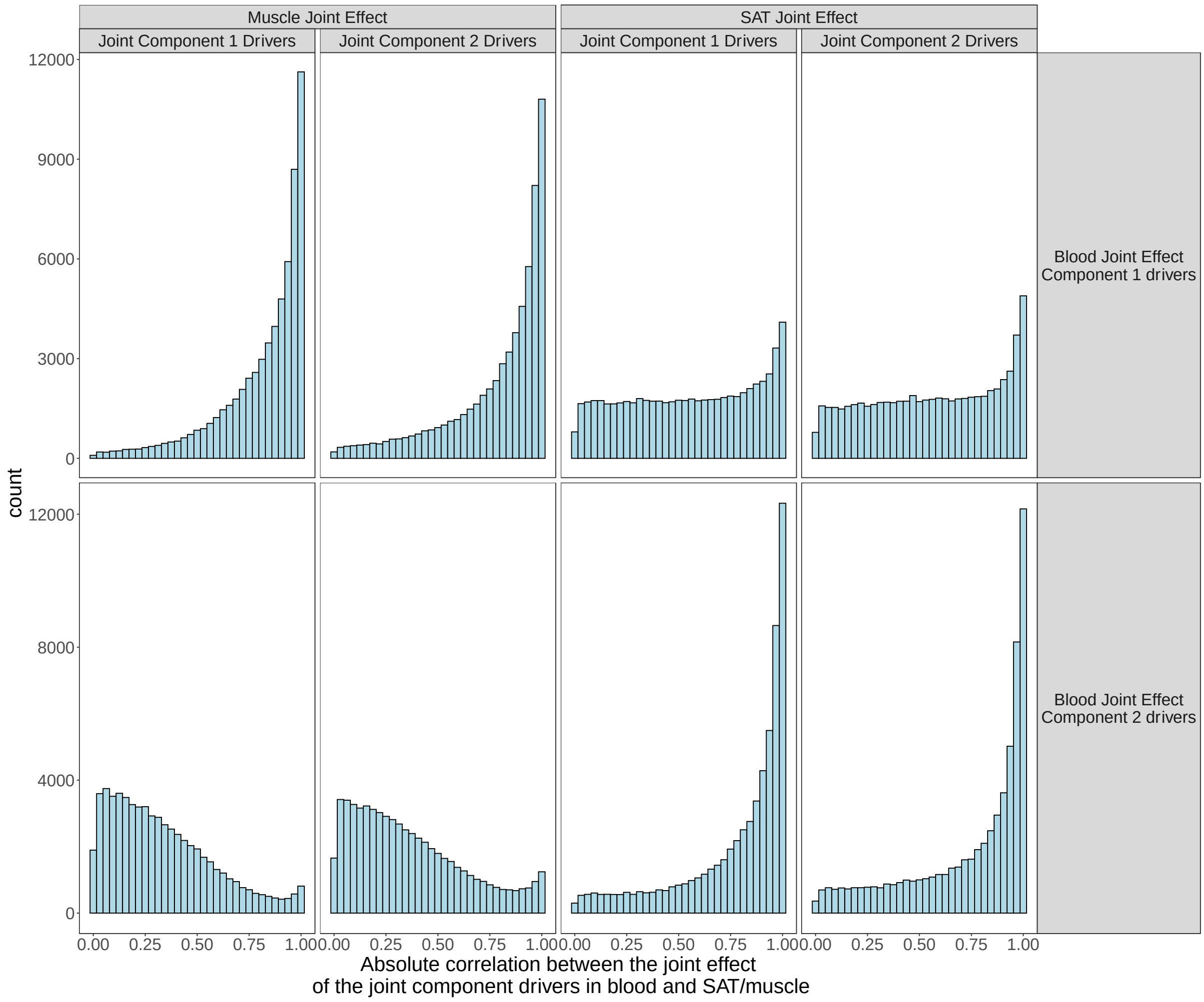


***Supplementary Figure 3: The drivers of the joint component 1 and 2 joint effect in blood are associated to the joint effect in muscle and SAT, respectively.*** *Histogram of the absolute correlations between the joint effect of the rank 1 and drivers of blood, and SAT and muscle. The distribution of the absolute correlation between the joint effect in blood and the other two tissues is plotted on the x-axis.*
